# Single cell multiomics reveals drivers of metabolic dysfunction-associated steatohepatitis

**DOI:** 10.1101/2025.05.09.25327043

**Authors:** Weston Elison, Lei Chang, Yang Xie, Charlene Miciano, Qian Yang, Hannah Mummey, Ryan Lancione, Sierra Corban, Sadatsugu Sakane, Jacinta Lucero, Sainath Mamde, Hyun Young Kim, Matthew J Kim, Rebecca Melton, Luca Tucciarone, Audrey Lie, Timothy Loe, Tanmayi Vashist, Kelsey Dang, Ruth Elgamal, Daofeng Li, Melissa Vu, Elie N Farah, Chad Seng, Jovina Djulamsah, Bing Yang, Justin Buchanan, Michael Miller, Mai Tran, Janeth Ochoa Birrueta, Neil C Chi, Ting Wang, Agnieszka D’Antonio-Chronowska, Allen Wang, Tatiana Kisseleva, David Brenner, Bing Ren, Kyle J Gaulton

## Abstract

Metabolic dysfunction-associated steatotic liver disease (MASLD) has limited treatments, and cell type-specific regulatory networks driving MASLD represent therapeutic avenues. We assayed five transcriptomic and epigenomic modalities in 2.4M cells from 86 livers across MASLD stages. Integrating modalities increased annotation of the genome in liver cell types several-fold over previous catalogs. We identified cell type regulatory networks of MASLD progression, including distinct hepatocyte networks driving MASL and mild and severe fibrosis MASH. Our single cell atlas annotated 88% of MASH-associated loci, including a third affecting hepatocyte regulation which we linked to distal target genes. Finally, we characterized hepatocyte heterogeneity, including MASH-enriched populations with altered repression, localization, and signaling. Overall, our results provide high-resolution maps of liver cell types and revealed novel targets for anti-MASH therapy.

## Main text

Metabolic dysfunction-associated steatotic liver disease (MASLD) and alcohol-associated liver disease (MetALD) are the major causes of liver fibrosis in patients with metabolic syndrome, which consists of obesity, insulin resistance, and hyperlipidemia (*1*, *2*). MASLD is a spectrum of liver disease ranging from steatosis (metabolic dysfunction-associated steatotic liver [MASL]) to metabolic dysfunction-associated steatohepatitis ([MASH]), characterized by the development of steatosis, inflammation and fibrosis (*2*). Excessive alcohol consumption often exacerbates liver injury in MASH patients. (*3*, *4*). Over a third of the world population is affected by MASLD (*5*, *6*), of which an estimated 20% have MASH, and although MASH causes increased morbidity and mortality, there are few therapeutic options (*2*, *7*, *8*).

Studies in humans and experimental models have proposed a two-hit model for MASH and MetALD progression (*2*, *9*). Chronic metabolic injury of hepatocytes causes steatosis, hepatocyte damage, apoptosis (*10*), and subsequent recruitment of inflammatory cells (*11–14*), which drive activation of hepatic stellate cells (aHSCs/myofibroblasts) that secrete Collagen Type I and extracellular matrix proteins and make the liver fibrotic (*2*, *7*). The crosstalk between hepatocytes (*15*, *16*), myeloid cells (*17*, *18*), HSCs (*19–23*), HSCs, and liver sinusoidal endothelial cells (LSECs) plays a critical role in the pathogenesis of MASH (*21*, *24–26*). Single cell transcriptomic studies have provided initial insights into gene expression changes in these cell types during MASLD (*16*, *27–30*). However, characterization of the gene regulatory programs and signaling responses within liver cell types that drive MASH progression, are critical for understanding of how to modulate cellular function to prevent or reverse metabolic liver disease.

The epigenome describes covalent modifications to DNA and histone proteins throughout chromosomes, which collectively encodes multiple layers of gene regulation that define cell type-specific gene activity, signaling responses, and function (*31*, *32*). Previous single cell chromatin accessibility studies have defined reference maps of cell type-specific candidate *cis*-regulatory elements (cCREs) in the liver but lack information on functional context of cCRE activity, diversity of cCRE activity across individuals and MASLD states, and annotation of the non-coding genome outside of cCREs defined from accessible chromatin. Recently developed single cell assays enable profiling additional modalities describing the epigenome in specific human cell types (*33*– *36*). In this study we leveraged these advances to comprehensively profile single cell transcription, accessible chromatin regions, two histone modifications associated with active (H3K27ac) and repressive (H3K27me3) states, and 3D chromatin conformation in 86 human livers, along with spatial transcriptomics in seven livers, spanning the spectrum of MASLD progression. We defined cell type-specific gene regulatory programs and chromatin state in the liver, determined transcriptional drivers of progression to MASH and annotated genetic risk loci for MASLD in liver cell types, and characterized heterogeneity including MASH-associated sub-clusters in hepatocytes. Overall, we identified novel genes, pathways and transcriptional networks that could serve as targets for therapeutic intervention in MASH.

## Results

### Single cell and spatial map of the human liver from non-disease and MASLD donors

We performed single cell assays to profile gene expression, accessible chromatin, two histone modifications, and 3D genome architecture in human livers from 86 donors classified as normal (n=26), simple steatosis (MASL) (n=25), metabolic dysfunction associated steatohepatitis (MASH) (n=23) and MASH with history of alcohol consumption (MetALD) (n=12) (**Supplementary Table 1**). The MASH/MetALD donors were further divided into livers with mild/low-fibrosis (Fib- MASH; stages 0-2, n=19) and severe/high-fibrosis (Fib+ MASH; stages 3-4, n=16). An overview of the study design and assays performed for each donor is shown in **Figure 1A** and **Supplementary** Figure 1.

**Figure 1.**
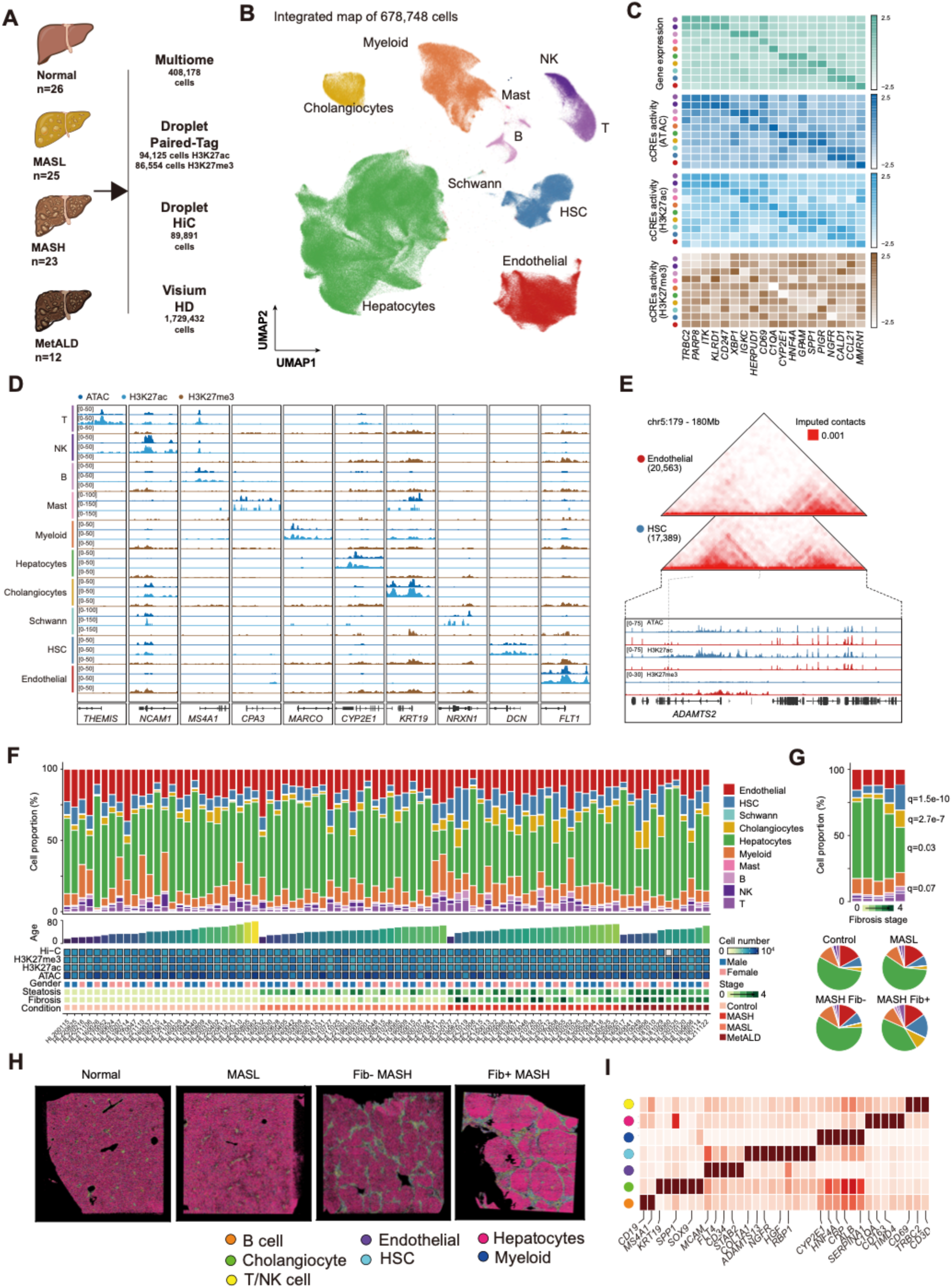
Integrated cell type-specific map of gene regulation in the human liver. (A) Study design consisting of profiling normal, MASL, MASH and MetALD livers from 86 human donors. Within MASH and MetALD livers were further classified by low fibrosis (Fib-) and high fibrosis (Fib+). Each donor was assayed using single cell multiome (paired ATAC-seq and RNA-seq), Paired tag for H3K27ac and H3K27me3 marks (paired Cut&Tag and RNA-seq), droplet Hi-C and genome-wide genotyping. A subset of donors was also assayed using spatial transcriptomics (Visium) (B) UMAP plot showing clustering of 678,748 nuclei from multiome, Paired-Tag and droplet Hi-C. Clusters are labeled based on cell type identity which is defined using gene expression of key marker genes. (C) Activity of selected cell type marker genes across liver cell types in gene expression, ATAC-seq, H3K27ac and H3K27me3 profiles. (D) Genome browser locus plots of selected cell type marker genes across liver cell types in ATAC-seq, H3K27ac and H3K27me3 profiles. (E) Chromatin contacts from Hi-C at the *ADAMTS2* locus in HSCs (blue) and endothelial (red) cells. Bottom shows the ATAC-seq, H3K27ac and H3K27me3 profiles in HSCs and endothelial cells for the same genomic region. (F) Liver cell type proportions (top), number of cells from each assay (middle), and MASLD phenotypes (bottom) across all 86 donors profiled in the study. (G) Proportions of each liver cell type across fibrosis stages (top) and MASLD state (bottom). (H) Cell type annotations in spatial imaging of representative normal, MASL, Fib- MASH and Fib+ MASH livers. We note that the colors denoting each cell type differ from the rest of the figure for the purposes of visualization of the spatial profiles. (I) Expression of selected marker genes of liver cell types in spatial profiles.

For all 86 liver donors, we first generated joint single nucleus accessible chromatin and gene expression (10x multiome) profiles using a pooled sample design, along with genome-wide genotyping (**Supplementary** Figure 1). After sample demultiplexing and removal of low-quality cells and doublets, we clustered 408,178 profiles across all 86 donors using single cell transcriptomes in Seurat (*37*) (**Supplementary** Figure 2). We annotated the cell type identity of multiome clusters using marker gene expression which identified hepatocytes, cholangiocytes, endothelial, myeloid, hepatic stellate cell (HSC), lymphoid, and other cell types (**Figure 1B,C, Supplementary Table 2**). Next, we generated joint single nucleus profiles of gene expression and histone modification for the active chromatin mark H3K27ac and repressive mark H3K27me3 using droplet-based Paired-Tag (*35*) on pooled samples from the same 86 donors (**Supplementary** Figure 1). Following demultiplexing and removal of low-quality cells and doublets, we reference mapped 94,125 H3K27ac and 86,554 H3K27me3 cells to the multiome atlas (**Supplementary** Figure 2). Finally, we performed droplet-based single cell Hi-C (*34*) on pooled samples from 85 of the 86 liver donors (**Supplementary** Figure 1). After demultiplexing and doublet removal from each pool, we obtained a total of 89,891 nuclei with high quality Hi-C profiles based on the number of total, *cis*, and *trans* chromatin contacts (**Supplementary** Figure 2). We integrated single-cell Hi-C profiles with the multiome and Paired-Tag map by using label transfer in Seurat (*38*) (**see Methods, Supplementary** Figure 2).

Overall, the resulting integrated map of the human liver consisted of a total of 678,748 single nuclei combining data from all multiome, Paired-Tag and Hi-C assays (**Figure 1B, Supplementary** Figure 2). Epigenomic profiles at cell type marker genes revealed accessibility (ATAC-seq) and active signal (H3K27ac) for the corresponding cell type and repressed signal (H3K27me3) for non-active cell types (**Figure 1C,D**). We also observed cell type-specific patterns of 3D chromatin architecture at known cell type marker genes, such as increased chromatin contacts at the *ADAMTS2* locus in HSCs compared to other liver cell types such as endothelial cells (**Figure 1E**). We further annotated single cell profiles at an additional layer of resolution to identify sub-types of cell types in the liver. This revealed known cell sub-types such as quiescent and activated HSCs (aHSCs), meschenchymal cells, sinusoidal endothelial cells (LSECs), liver resident Kupffer cells (KCs), myeloid cells (including lipid-associated macrophages) and other inflammatory cells (e.g. T/B/NK cells), and zone-specific hepatocytes, among other sub-types (**Supplementary** Figure 3).

We next examined the distribution of cell types and cell sub-types within each of the 86 liver donors. Each donor contributed consistently to the single-cell profiles across both multiome and Paired-Tag data, with balanced representation across phenotypes such as age, sex, and BMI (**Figure 1F**). Similarly, single cell Hi-C profiles showed consistent representation across donors (**Figure 1F**). We observed significant changes (FDR<.10) in the abundance of liver cell types across MASLD states, most prominently an increased abundance of HSCs in MASH, particularly in high-fibrosis (Fib+) MASH, reflecting their activation in MASLD (**Figure 1F,G**). In addition, cholangiocytes, B cells and T cells were all increased, and hepatocytes decreased, in MASH progression, consistent with increased inflammation and hepatocyte loss (**Figure 1F,G, Supplementary Table 3**). There were similarly significant changes (FDR<.10) in the abundance of cell types in higher fibrosis stage including increased HSCs, cholangiocytes, and B cells, as well as decreased hepatocytes (**Figure 1F,G, Supplementary Table 3**). We also observed significant changes (FDR<.10) in the abundance of liver cell sub-types in MASLD; for example, aHSCs, vascular and lymphatic endothelial cells, and lipid-associated macrophages were all more abundant across MASH progression and in higher fibrosis stage (**Supplementary Table 3**).

Finally, we used spatial transcriptomics (Visium HD) to profile the organization of liver cell types in seven livers representing normal (n=2), MASL (n=2), MASH (n=2) and MetALD (n=1) (**see Methods, Supplementary** Figure 4). After cell segmentation and filtering of low quality cells, we annotated 1,729,432 cells based on reference mapping from the single cell multiome map using Tangram (*39*). Cell type assignments were confirmed based on expression of key marker genes (**Figure 1H,I**). Normal livers were composed of liver lobules with central and portal veins and hepatocytes organized in cords, while quiescent HSCs resided in the space of Disse and myeloid cells were found throughout liver parenchyma. In MASL, normal liver architecture was preserved but with steatosis, characterized by accumulation of lipid droplets in hepatocytes. MASH livers had injured hepatocytes around the central vein, inflammation, and fibrosis. Low fibrosis (Fib-) MASH was characterized by aHSCs with migration towards liver injury and inflammation. High fibrosis (Fib+) MASH or cirrhosis had severe inflammation with bridging fibrosis, and aHSCs, myeloid, T and B cells near the fibrous scar surrounding hepatic regenerative nodules.

Together, these results provide an integrated single cell map and spatial profiles from normal and MASLD livers. To provide these genomic data to the community as a resource, we developed a browser available at https://epigenome.wustl.edu/MASLD/.

### Cell type-specific gene regulatory programs in the liver

We next leveraged the integrated single cell map of multiple modalities to characterize the gene regulatory programs of each liver cell type. We first defined a unified set of 611,576 candidate *cis*-regulatory elements (cCREs) across cell types using accessible chromatin (ATAC-seq) profiles (**Supplementary Data**). Compared to previous catalogs of cCREs in the adult and fetal liver (*32*, *40*), between 43.50-47.89% of cCREs we annotated were novel (**Figure 2A, Supplementary** Figure 5). Liver cell type cCREs overall showed expected distribution across genomic features, although novel cCREs were more enriched in retro-transposon and other classes of elements relative to known cCREs (**Figure 2A, Supplementary** Figure 5). The novel cCREs also exhibited greater inter-donor variability than known cCREs, consistent with the expanded number and diversity of profiled livers uncovering a larger portfolio of regulatory elements (**Supplementary** Figure 5).

**Figure 2.**
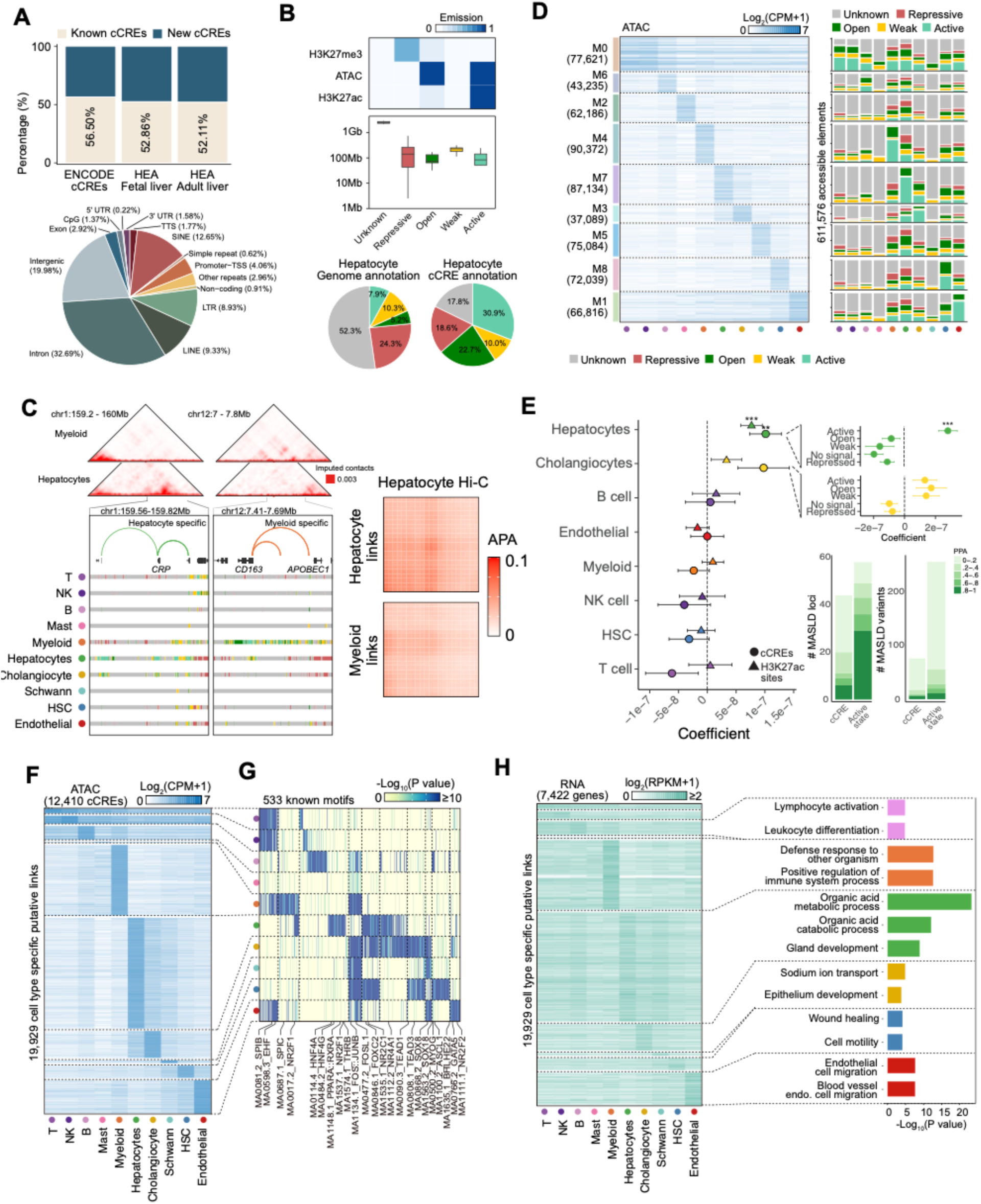
Cell type-specific gene regulatory programs in the human liver. (A) Percentage of liver cell type cCREs in this study identified in the liver in ENCODE and HEA (top), and distribution of liver cell type cCREs across various genomic features (B) Emission probabilities of ATAC-seq, H3K27ac and H3K27me3 for each chromatin state (active, open, weak, repressive, and unknown) and proportion of the genome annotated with each state in each cell type (top), and distribution of chromatin states in hepatocytes genome-wide (bottom left) and at cCREs (bottom right). (C) Examples of gene regulatory networks (GRNs) with hepatocyte-specific and myeloid-specific cCRE-target gene links and active chromatin state at the *CRP* and *CD163* loci, respectively. Chromatin contacts in hepatocytes and myeloid cells support cell type-specific activity at each locus. Hepatocyte cCRE-target gene links from GRNs show enrichment of chromatin contacts compared to proximal regions, while myeloid target genes links show no enrichment. (D) Clustering of liver cell type cCREs into modules (M0-8) based on accessibility profiles across cell types (left) and chromatin state of cCREs in each module across cell types (right). (E) Enrichment of chronic alanine aminotransferase (cALT) level associated variants in cCREs and H3K27ac sites in each liver cell type (left), enrichment of cALT associated variants in cCREs annotated in each chromatin state in hepatocytes and cholangiocyte (top right), and number of fine-mapped cALT loci and variants overlapping hepatocyte cCREs and active chromatin states. Colors represent the posterior probability (PPA) of variants. (F) Liver cell type cCREs with accessibility profiles highly specific to a cell type. Plotted values represent log scaled counts-per-million (CPM) in the cCRE. (G) Transcription factor sequence motifs enriched in cCREs with cell type-specific accessibility for each cell type. (H) Expression level of genes in each cell type linked to cCREs with cell type-specific accessibility (left), and biological pathways enriched in genes linked to specific cCREs in each cell type (right).

Next, we called chromatin states in each cell type using ATAC-seq, H3K27ac, and H3K27me3 profiles with chromHMM (*41*), which revealed active (ATAC-seq+H3K27ac), open (ATAC-seq), repressive (H3K27me3) and weakly active chromatin states (**Figure 2B, Supplementary Data**). Chromatin states annotated a substantially larger proportion of the non-coding genome in liver cell types compared to cCRE definitions. For example, in hepatocytes, 47.7% of the genome was annotated as an active, weak, open or repressed chromatin state, compared to 5.9% of the genome in cCREs based on chromatin accessibility only (**Figure 2B**). Comparing chromatin states in liver cell types to states in bulk liver from NIH Epigenome Roadmap (*42*) revealed strong concordance in state identity across the genome for hepatocytes with lower concordance among less common cell types (e.g. HSCs) (**Supplementary** Figure 5), highlighting that bulk chromatin state profiles more closely reflect common cell types. As the functional roles of most cCREs are undefined, we used chromatin state information to annotate cCREs in each cell type. In hepatocytes, for example, 30.9% of all cCREs were in an active state while 10% and 22.7% were weak and open states, respectively, and 36.4% were repressed or unknown (**Figure 2B**). The cCREs annotated in different chromatin states had distinct properties; for example, in hepatocytes, cCREs in active states were enriched for HNF1/4, FOXA and PPAR TF motifs while cCREs in open states were enriched for CTCF, CUX1/2 and ONECUT motifs and cCREs in repressed states were enriched for REST motifs (**Supplementary** Figure 5).

We defined TF gene regulatory networks (GRNs) in each cell type using SCENIC+ (*43*). Cell types had GRNs for 186 TFs (105-137 per cell type) and, on average, TF GRNs consisted of 123 cCREs linked to 82 genes (**Supplementary Data**). Biological pathways enriched in target genes of GRNs revealed known transcriptional regulators of cellular processes - for example, in hepatocytes, the STAT1 GRN was linked to interferon signaling and viral response and the ATF3/4 GRN was linked to endoplasmic reticulum stress and unfolded protein response - as well as many putative relationships (**Supplementary Table 4**). We utilized 3D chromatin contacts in each cell type to validate target gene predictions of cCREs in GRNs. For example, at the *CRP* locus, multiple hepatocyte-specific cCRE links were supported by chromatin contacts in hepatocytes and not in myeloid cells (**Figure 2C**). Similarly, at the *CD163* locus, myeloid-specific cCRE links were supported by chromatin contacts in myeloid cells (**Figure 2C**). More broadly, cCRE-target gene links collectively had increased 3D chromatin contacts for the corresponding cell type compared to other cell types (**Figure 2C**). We also identified physical interactions between distal regions by calling chromatin loops using the 3D chromatin contacts in liver cell types with sufficient coverage (hepatocytes, endothelial, HSC, myeloid cells). Hepatocytes had the largest number of detected loops with fewer loops in less abundant cell types (**Supplementary Data**).

We next examined the chromatin states of cCREs across liver cell types to determine how their activity varies across cell types. We grouped all 611,576 cCREs into modules based on accessible chromatin signal across cell types, revealing nine distinct modules (M0-M8) (**Figure 2D**). One module (M0) had broadly shared accessibility across cell types while other modules had accessibility profiles that were more cell type restricted (**Figure 2D**). We next determined the chromatin state of cCREs in each module. The shared (M0) module was broadly enriched for active chromatin states across cell types, and a contained higher proportion of cCREs in the M0 module mapping to gene promoters relative to other modules (13.4% vs. 2.5%). By comparison, the other cell-type restricted modules were enriched for active and open chromatin states in the corresponding cell type(s) where accessibility was observed (**Figure 2D**).

The liver cell types and regulatory programs through which genetic variants influence MASLD risk remain poorly defined. We utilized genome-wide association study (GWAS) data for circulating alanine aminotransferase (cALT) level (*44*), a proxy for liver damage and MASLD, to identify genetic determinants of MASLD in liver cell types. We identified significant enrichment (FDR<.10) of MASLD-associated variants genome-wide in hepatocyte cCREs using LD score regression (*45*, *46*), as well as a similar magnitude of enrichment in hepatocyte H3K27ac sites (**Figure 2E**). When further classifying cCREs by chromatin state, we observed stronger enrichment of MASLD variants for hepatocyte cCREs in active states and no corresponding enrichment for other states (**Figure 2E**). We also observed significant enrichment of MASLD variants among cCREs in the hepatocyte-specific module (M7) and no corresponding enrichment for other modules (**Supplementary** Figure 6).

We next annotated specific MASLD-associated loci where, overall, 348 out of 659 fine-mapped variants with PPA>.01, representing 68/77 (88%) all MASLD loci, were annotated to liver cell type cCREs or active states (**Supplementary Table 5, Supplementary** Figure 7). When considering just hepatocytes, 274 fine-mapped variants with PPA>.01 at 58 loci were annotated to hepatocyte cCREs or active states, where active states annotated a larger number of variants compared to cCREs (**Figure 2E**). Among variants in hepatocyte cCREs or active chromatin states were several with high causal probabilities (PPA>.99) and unannotated in previous catalogs of liver cell type cCREs (*40*), such as rs4841132 at the *PPP1R3B* locus and rs11621792 at the *NFATC4* locus.

Outside of hepatocytes, several loci were annotated specifically in other cell types such as *TRPS1* (indexed by rs2737217) and *HLA-DQA1* (rs686250) in myeloid cells, *FCGR2B* (rs74816838) in endothelial cells, and *FLACC1* (rs10201587) in HSCs, supporting that these cell types also play a role in MASLD risk (**Supplementary** Figure 7).

Finally, we identified gene regulatory programs that specify the identity of each liver cell type. From the set of 611,576 cCREs, we identified 32,663 (5.3%) cCREs with cell type-specific accessibility profiles, of which 12,410 of these cCREs were linked to a putative target gene (**Figure 2F, Supplementary Table 6**). Sequence motifs enriched in cell type-specific cCREs revealed key regulators of cell type identity such as HNF4A, RXRA and NR2F1 TF motifs for hepatocytes, SPIB and SPIC motifs for myeloid cells, SOX and GATA motifs for endothelial cells, and FOS, JUN and TEAD motifs for HSCs (**Figure 2G**). Cell type-specific cCREs in hepatocytes were significantly and specifically enriched for cALT level associated variants, confirming the prominent role of hepatocytes in MASLD risk (**Supplementary** Figure 6). Cell type-specific cCREs were linked to 7,422 target genes through GRNs, and these target genes overall showed corresponding cell type-specific expression (**Figure 2H, Supplementary Table 6**). Furthermore, cell type-specific target genes were enriched for key process related to cellular function and identity, such as cholesterol transport and glycine metabolism in hepatocytes (**Figure 2H**).

Together, these results provide an expanded annotation of gene regulatory programs of cell types in the human liver which facilitates annotation of variants involved in MASLD risk.

### Changes in liver cell type gene regulation associated with MASLD

Given a comprehensive definition of gene regulatory programs in liver cell types, we next determined how cell type regulatory programs change across progression to MASH. We observed marked changes (FDR<.10) in gene expression, chromatin accessibility, and histone modification profiles across MASLD stages compared to normal livers in hepatocytes, cholangiocytes, HSC, endothelial, and myeloid cells (**Figure 3A, Supplementary** Figure 8**, Supplementary Data**). In each cell type, the number of gene or epigenomic features with significant changes in activity compared to normal livers was substantially more pronounced in MASH and MetALD than for MASL (**Figure 3A**). Moreover, within MASH/MetALD livers, significant changes were more extensive in high fibrosis (Fib+) MASH compared to low fibrosis (Fib-) MASH (**Figure 3A, Supplementary** Figure 8**, Supplementary Data**), highlighting fibrosis as a major cause of disease-associated changes.

**Figure 3.**
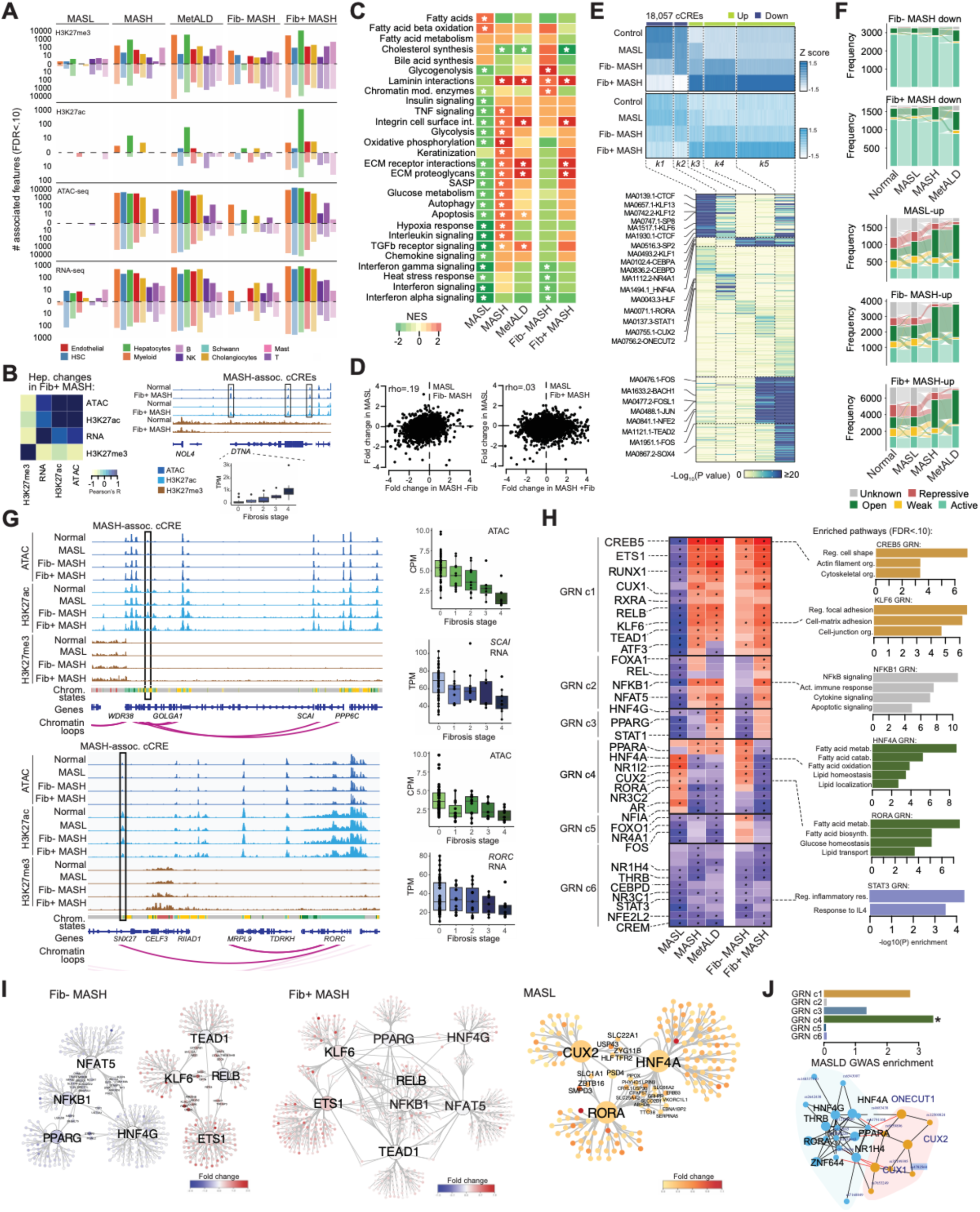
Cell type-specific genomic and epigenomic changes in MASLD. (A) Number of genomic features in each cell type for each modality with significant changes in activity in different MASLD stages compared to normal livers. (B) Pearson correlation in fold-change in hepatocyte activity in high fibrosis (Fib+) MASH across modalities (left), and genome browser plot of the *DTNA* locus where, in Fib+ MASH, several cCREs had increased accessibility and H3K27ac signal, *DTNA* had increased expression, and the region had broadly reduced H3K27me3 signal. (C) Biological pathways in hepatocytes with altered expression in MASLD stages compared to normal livers. Values shown in the heatmap are normalized enrichment scores (NES) and cells contain a * if the NES were significant at FDR<.10 (top), and correlation in the fold-change of gene expression levels in hepatocytes in MASL compared to Fib- and Fib+ MASH (bottom). (D) Clusters of hepatocyte cCREs with significant change in Fib+ MASH across different MASLD stages in ATAC-seq, H3K27ac and H3K27me3 profiles (top), and transcription factor sequence motifs enrichments for cCREs in each cluster (bottom). (E) Chromatin state in each MASLD stage for hepatocyte cCREs with decreased activity in MASH (top) and increased activity in MASL/MASH (bottom). (F) Genome browser tracks showing example loci with altered activity in hepatocytes in MASH. A hepatocyte cCRE in the *GOLGA1* gene (highlighted in black rectangle) with decreased activity in higher fibrosis and in an active hepatocyte chromatin state shows physical interactions via chromatin looping with the promoter region of the *SCAI* gene, which has decreased expression higher fibrosis (top); A hepatocyte cCRE in the *SNX27* gene (highlighted in black rectangle) with decreased activity in higher fibrosis and in an active hepatocyte chromatin state shows physical interactions via chromatin looping with the promoter region of the *RORC* gene, which has decreased expression higher fibrosis (bottom). (G) Transcription factor gene regulatory networks (GRNs) in hepatocytes with increased or decreased activity in MASLD stages compared to normal livers. GRNs are grouped (GRN c1-6) based on change in activity across MASLD stages. Cells with significant (FDR<.10) change in GRN activity are highlighted with * (left), and biological pathways significantly (FDR<.10) enriched among genes in each GRN (right). (H) Connectivity of GRNs with altered activity in Fib- MASH, Fib+ MASH and MASL. Transcription factor genes as well as the genes in GRNs for each factor are colored based on the change in expression in the specified MASLD stage. (I) Fold-enrichment of fine-mapped MASLD (cATL) associated variants for hepatocyte GRNs in each GRN group compared to background cCREs (top), and connectivity of hepatocyte GRNs based on fine-mapped MASLD variants overlapping a cCRE in the GRN. Colors are based on the clustering of GRN connectivity using igraph.

Notably, we observed a larger number of significant H3K27ac and H3K27me3 features with altered activity in hepatocytes of Fib+ MASH livers compared to other cell types (**Figure 3A, Supplementary Data**). We determined the relationship between changes in genomic modalities in hepatocytes in Fib+ MASH livers, which revealed overall positive correlation in changes in gene expression, accessible chromatin and H3K27ac activity and a negative correlation with the repressive mark H3K27me3 (**Figure 3B**). For example, across the *DTNA* locus there was a loss of repression in hepatocytes in Fib+ MASH, which coincided with increased cCRE activity and the expression of *DTLA* (**Figure 3B**). For example, across the *DTNA* locus there was a loss of repression in hepatocytes in Fib+ MASH, which coincided with increased hepatocyte cCRE activity and the expression of *DTNA*, a component of the dystrophin-associated protein complex (DPC) that binds cytoskeleton and extracellular matrix (**Figurer 3B**).

We identified pathways enriched among genes with altered expression in MASH compared to normal livers (**Figure 3C, Supplementary Table 7**). Hepatocytes in MASH had increased expression of pathways related to epithelial remodeling, such as cytoskeletal structure and cell-ECM interactions, although they did not contribute to deposition of fibrillar collagens or ECM (**Supplementary** Figure 9**, Supplementary Table 7**), as well as pathways related to glycolysis, hypoxia, senescence, cytokine signaling, cell death, and signaling responses (**Figure 3C, Supplementary Table 7**). In turn, MASH hepatocytes exhibited reduced expression of genes in fatty acid and cholesterol related pathways. In MASH livers, HSCs upregulated Hedgehog signaling, Ras signaling, fiber formation, collagen degradation, and matrix metallothionein activity, while endothelial cells had increased ECM interactions and decreased inflammatory signaling (**Supplementary Table 7**). When further characterizing changes in hepatocytes in Fib+ and Fib- MASH livers, the strong enrichment in cytoskeletal and cell-ECM interaction pathways (**Figure 3C**) observed in MASH livers could be attributed to Fib+ MASH, while these pathways had either less pronounced induction or no effect in Fib- MASH hepatocytes (**Figure 3C**). In turn, Fib- MASH livers showed distinct up-regulation of fatty acid metabolism, synthesis, and oxidation pathways compared to normal livers.

As both MASH and MetALD are caused by metabolic liver injury with exacerbated hepatic inflammation and fibrosis due to chronic alcohol consumption in the latter, we next assessed their molecular differences and similarities. Despite profiling fewer MetALD livers, we identified more genes and cCREs overall with significant changes in activity in MetALD compared to MASH (**Figure 3A, Supplementary Data**). Changes in individual gene and pathway expression in MASH and MetALD livers were highly correlated across cell types (all r^2^>.5, P<2x10^-16^), although gene effects were overall more pronounced in MetALD (hepatocytes binomial p=2x10^-16^). Among specific pathways, MetALD hepatocytes had more pronounced activity of cell-ECM interaction pathways while MASH hepatocytes showed increased interleukin signaling, glycolysis, oxidative phosphorylation, and respiration. In MetALD HSCs, there was increased cell-ECM interaction pathways, while MASH HSCs had increased oxidative phosphorylation, renin-angiotensin system (RAS) processing, translation and metabolism pathways (**Supplementary Table 7**).

While MASL showed few significant changes in expression of individual genes compared to normal livers, numerous pathways were broadly altered in MASL in hepatocytes and other cell types (FDR<.10) (**Figure 3C, Supplementary** Figure 8**, Supplementary Table 7**). There was significant correlation in effects on gene expression in MASL and MASH in non-parenchymal cells (including HSCs, endothelial cells, and cholangiocytes) (rho>.3), yet hepatocytes showed little to no correlation (rho=.05), highlighting the distinct gene-regulatory programs of MASL and MASH hepatocytes. When separating MASH into Fib+ and Fib- livers, there was a modest correlation in effects between MASL and Fib- MASH (rho=.19) and very low correlation with Fib+ MASH (rho=.03) (**Figure 3D**). Further, MASL and MASH exhibited negative correlation (rho=-.30) in pathway enrichments compared to normal livers. In contrast to MASH, hepatocytes in MASL up- regulated fatty acid metabolism and biosynthesis and did not show increased epithelial remodeling, inflammation, and fibrotic signaling (**Figure 3C, Supplementary Table 7**). Overall, this supports the concept that MASL and MASH are driven by divergent gene expression programs in hepatocytes. In Fib− MASH livers, hepatocytes retained characteristics of MASL hepatocytes while also sharing some features with Fib+ MASH hepatocytes (**Figure 3C**), suggesting it may represent a transitional state between MASL and advanced fibrosis MASH.

Next, we examined epigenomic changes in hepatocytes associated with progression to MASH. There were 18,057 and 49,422 cCREs with significant (FDR<.10) changes in activity in Fib+ MASH and fibrosis score, respectively, where these cCREs had concordant changes in both chromatin accessibility and H3K27ac signal (**Figure 3E, Supplementary** Figure 10). Clustering of MASH-associated cCREs revealed multiple sets of up-regulated cCREs, including those altered starting in MASL and others altered only in MASH, as well as down-regulated cCREs (**Figure 3E, Supplementary** Figure 10). To further understand cCRE dynamics, we defined chromatin states separately in MASLD stages and categorized cCREs by state across progression to MASH. Hepatocyte cCREs with increased and decreased accessibility in higher fibrosis score livers exhibited a progressive gain and loss of active chromatin states, respectively, as well as, in the former, loss of repressive states (**Figure 3F, Supplementary** Figure 11). We also observed chromatin state changes in hepatocytes from normal to MASL livers, including subsets of cCREs in open states in normal livers that transitioned to active states in MASL and vice versa (**Supplementary** Figure 11). We next identified the gene targets of MASH-associated cCREs using chromatin loops and TF GRNs in hepatocytes (**Figure 3G, Supplementary Table 8**). For example, distal cCREs at the 9q33 and 1q21 loci with decreased hepatocyte accessibility and H3K27ac activity in Fib+ MASH interacted with the promoter region of *SCAI* and *RORC,* respectively, both of which had reduced expression in MASH **(Figure 3G**), and where the latter is involved in insulin signaling and lipid metabolism.

To identify upstream regulators of cCRE activity in MASLD, we identified TF sequence motifs enriched in cCREs with altered activity in MASLD. In hepatocytes, there were distinct motif enrichments among the different clusters of cCREs with altered activity in Fib+ MASH and high fibrosis score (**Figure 3E, Supplementary Table 9,10**). For example, cCREs with up-regulated activity starting in MASL were enriched for HNF4A, NR4A1, CEBPA/D TF motifs. Similar motifs including NR4A1/2 and CEBP were also enriched in cCREs that transitioned to active chromatin states in MASL from normal livers while, conversely, MAF and CTCF motifs were enriched in cCREs that transitioned from active to open states in MASL (**Supplementary** Figure 11). In comparison, cCREs up-regulated only in MASH and MetALD were enriched in FOS/JUN, TEAD and NFKB motifs, and cCREs down-regulated in MASH were enriched for CTCF, KLF, SP, ELK and ETV motifs (**Figure 3E**). Together these findings further support that MASL and MASH in hepatocytes are characterized by distinct TFs and regulatory programs. We also observed TF motifs enriched in cCREs altered in other cell types. In MASH HSCs, cCREs with increased accessibility were enriched for TEAD, RUNX, FOS/JUN, ATF3 and hormone receptor motifs (GRE, ARE), previously implicated in HSC activation (*47*, *48*), while cCREs with decreased accessibility were enriched for farnesoid X receptor (FXR) and ETS/ETV motifs, associated with maintenance of quiescence in HSCs (*49*, *50*) (**Supplementary Table 9**). In MASH endothelial cells, cCREs with increased activity were enriched for FOS/JUN, TCF, and MEF motifs, while cCREs with decreased activity were enriched for IRF motifs (**Supplementary Table 9**).

We next sought to further define transcriptional drivers of progression to MASH in hepatocytes by identifying TF GRNs with altered activity across MASLD states. TF GRNs with increased activity in MASL included HNF4A, RORA, and CUX2 (**Figure 3H,I, Supplementary Table 11**). These MASL-enriched GRNs were highly connected to genes with increased expression in MASL and involved in lipid biosynthesis (*SMPD3, LPIN3, SLC25A42*), binding (*PSD4*), transport (*SLC22A1*), and metabolism (*CRYL1*) (**Figure 3I**). We also identified GRNs with increased activity in MASH, which were highly distinct from those in MASL, including CREB5, NFKB1/RELB, KLF6, TEAD1, ETS1, and NFAT5 (**Supplementary Table 11**). MASH GRNs each regulated genes with increased expression in MASH including those involved in cytoskeletal organization, adhesion and ECM interactions (*DTNA, AKAP12, ITGA2, LAMA3, MYO7B, CD44*), inflammation (*TNFAIP2*), autophagy (*TMEM150B*), and lipid metabolism (*FGF1*) (**Figure 3H,I**). The overall activity of TF GRNs was similar in MASH and MetALD, although MetALD showed relatively higher activity of HNF4G, STAT1, and PPARG GRNs (**Figure 3H, Supplementary Table 11**).

Stratifying MASH livers into Fib+ and Fib- groups revealed additional dynamics of TF activity and their associated gene networks (**Figure 3H,I**). For example, MASL hepatocytes uniquely induced HNF4A, NR1I2, CUX2, and RORA GRNs, while Fib- MASH hepatocytes shared some of these GRNs, uniquely upregulated FOXO1, NR4A1, and FOS, and activated GRNs characteristic of Fib+ MASH hepatocytes such as CREB5, ETS1, RUNX1, CUX1, RELB, KLF6, TEAD1, and ATF3. In turn, Fib+ MASH hepatocytes uniquely upregulated HNF4G, PPARG, NFKB, NFAT5 and STAT1 and downregulated PPARA and HNF4A GRNs, highlighting fibrosis as a primary driver of regulatory changes in MASH. We further grouped GRNs by activity across MASLD stages which revealed distinct clusters (c1-c5) of GRNs with increased activity in MASL and Fib- MASH, Fib-/Fib+ MASH, Fib- MASH only, and Fib+ MASH only (**Figure 3H**). Genes in GRNs with altered activity in MASL and MASH were enriched (FDR<.10) in pathways with corresponding changes in MASL and MASH expression, highlighting drivers of altered pathway activity in MASLD (**Figure 3H, Supplementary Table 3**). For example, GRNs with increased activity in MASL and Fib- MASH, such as HNF4A and RORA, were associated with regulation of lipid metabolism/synthesis; GRNs active in both Fib- and Fib+ MASH, including KLF6, ETS1, and CREB5, regulated epithelial remodeling processes such as cytoskeletal organization and cell-ECM adhesion; and GRNs with highest activity in Fib+ MASH, such as NFKB and RELB, regulated inflammation and immune signaling.

Finally, to understand genetic risk in GRNs associated with progression to MASH, we tested for enrichment of MASLD (cALT) associated variants in hepatocyte GRNs. In hepatocytes, variants showed enrichment for GRN clusters with increased activity in MASL and Fib- MASH (GRN c4) and with increased activity in both Fib- and Fib+ MASH (GRN c1), with limited enrichment in other GRN clusters (**Figure 3J**, **Supplementary** Figure 12). Specific TF GRNs annotated with MASLD variants included those increased in MASL and Fib- MASH (e.g. HNF4A, RORA, CUX2), increased in Fib- and Fib+ MASH (ETS, KLF6, ATF3), and decreased across all MASLD stages (THRB, ONECUT1). We identified multiple clusters of TF GRNs altered in MASL and Fib- MASH based on overlapping MASLD variants (**Figure 3J**), and these GRNs were linked to multiple genes (*MYCLD, MTTP, APOL2, MLXIP)* in pathways such as lipid metabolism and transport.

Overall, these results identify transcriptional drivers of progression to MASH in liver cell types and show that hepatocytes exhibit distinct gene regulatory programs between MASL and MASH with additional genomic changes associated with severe fibrosis in MASH.

### MASLD-associated variants affecting hepatocyte regulatory programs

We next sought to identify specific loci and genes affecting MASLD risk in hepatocytes by combining quantitative trait locus (QTL) mapping, variant functional predictions, and 3D chromatin architecture.

We performed QTL mapping of chromatin accessibility (caQTLs), histone modifications (hQTLs), and gene expression (eQTLs) levels in liver cell types across all 86 donors using TensorQTL (*51*) (**Supplementary Data**). In total, we identified significant QTLs (FDR<.10) for each modality, including 10,591 caQTLs, 1,644 eQTLs, 418 H3K27ac hQTLs, and 391 H3K27me3 hQTLs across all cell types (**Figure 4A, Supplemental Table 12, Supplementary** Figure 13). Across modalities, most QTLs were identified in hepatocytes, with fewer in less common cell types (**Figure 4A**). Cell type caQTLs and hQTLs disrupted distinct TF sequence motifs, including HNF, FOX, PPAR, CUX, RXR, and CEBP motifs in hepatocytes, CEBP, STAT, IRF, SPIC, and ETV TFs in myeloid cells, FEV, FLI1, ETS and KLF motifs in endothelial cells, and ETS and FOS motifs in HSCs (**Figure 4B, Supplemental Table 13**). Overall, caQTLs and hQTLs had highly cell type-specific effects compared to eQTLs which were more often shared across cell types (**Figure 4C**). Furthermore, caQTLs/hQTLs and eQTLs at the same locus were largely distinct based on statistical colocalization (*52*), suggesting they captured broadly different classes of variant regulatory effects (**Figure 4C**).

**Figure 4.**
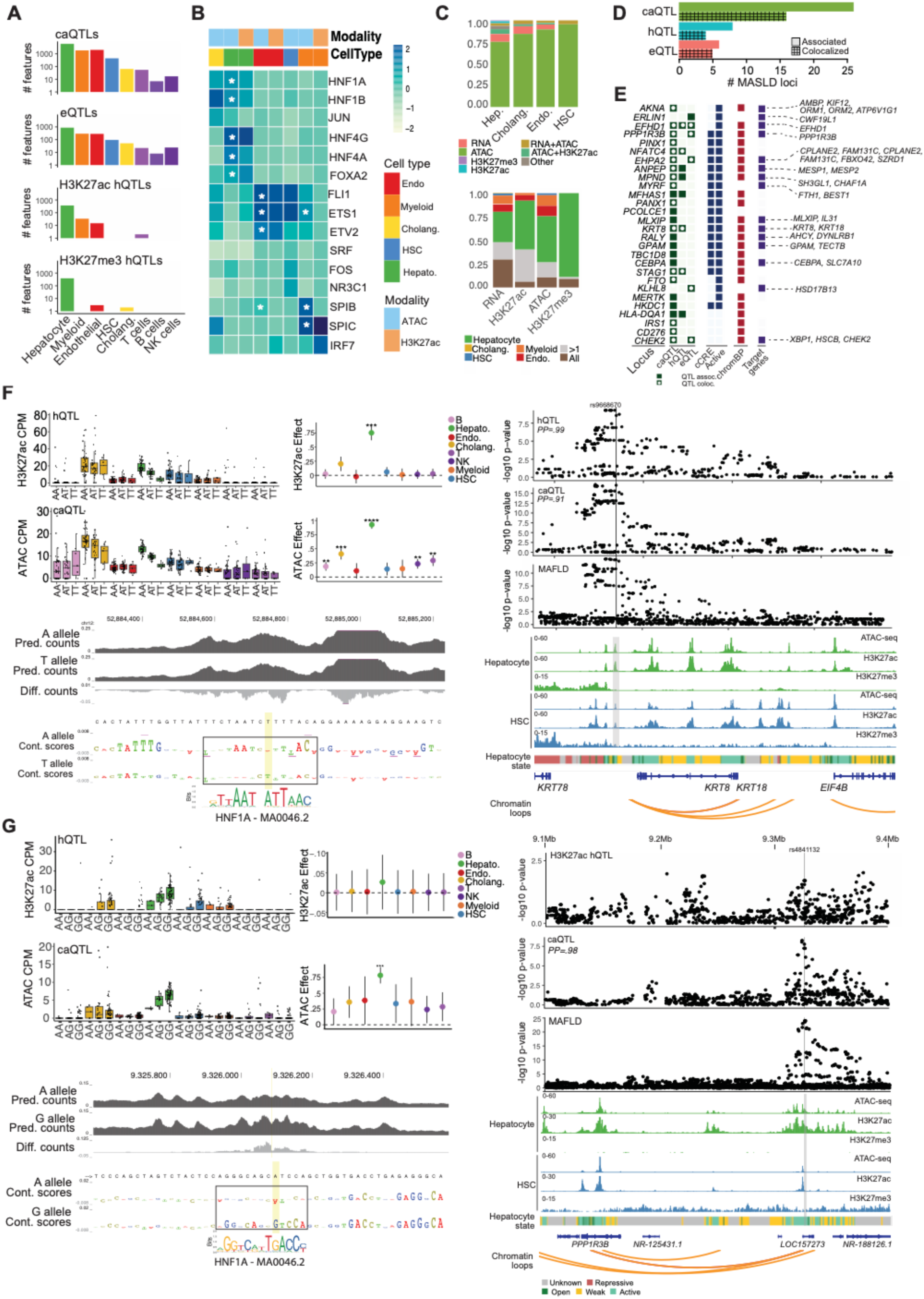
MASLD risk loci affect hepatocyte gene regulatory programs. (A) Number of features for each modality with a significant (FDR<.10) QTL in each liver cell type. (B) Transcription factor sequence motifs enriched in chromatin and H3K27ac QTLs for each cell type. *FDR<.10. (C) Proportion of QTLs in each cell type for each modality which are colocalized with QTLs for another modality or not colocalized with any other QTLs (top), and proportion of QTLs in each modality that are shared across cell types or show cell type specificity. (D) Number of MASLD-associated loci that are significant QTLs for each modality and colocalize with QTLs for each modality in hepatocytes. (E) Annotation of specific MASLD loci with QTL association including loci with a fine-mapped variant overlapping a cCRE or active chromatin state, a fine-mapped variant with predicted effects on hepatocyte chromatin with chromBPnet, and a link to a predicted target gene. (F) MASLD-associated variants at the 12q13 locus affect activity of a cCRE in hepatocytes, where the risk allele A has increased chromatin accessibility and H3K27ac activity (top left) and has higher predicted accessibility and maps in a sequence motif for HNF1A (bottom left). The MASLD association signal was strongly colocalized with chromatin accessibility and H3K27ac QTL association for the cCRE (highlighted in grey) in hepatocytes, and the cCRE region showed physical interactions in hepatocytes with the *KRT8* and *KRT18* promoter regions (right). (G) MASLD-associated variants at the 8p23 locus affect activity of a cCRE in hepatocytes, where the risk allele A has decreased chromatin accessibility and H3K27ac activity (top left) and has lower predicted accessibility and maps in a sequence motif for HNF1A (bottom left). The MASLD association signal was strongly colocalized with chromatin accessibility QTL association for the cCRE (highlighted in grey) in hepatocytes, and the cCRE region showed physical interactions in hepatocytes with the *PPP1R3B* promoter regions (right).

We next annotated MASLD-associated loci that affect hepatocyte regulatory programs. For these analyses we again utilized genetic association data for cALT levels, a proxy for liver damage and MASLD, due to the large number of established loci. Variants at over a third of all known cALT (MASLD) loci (28/77) were QTLs in hepatocytes, of which 18 were colocalized with QTL associated variants (**Figure 4D, Supplementary Table 14**). The majority of MASLD loci with QTLs (26/28) were caQTLs, including 11 and 15 loci where risk alleles increased and decreased cCRE activity, respectively (**Figure 4D**). Next, we prioritized target genes of loci affecting hepatocyte cCRE activity using eQTLs. Several MASLD loci colocalized with eQTLs, for example at the 2q37 locus where risk alleles increased cCRE accessibility, H3K27ac signal, and *EFHD1* expression in hepatocytes (**Figure 4D,E, Supplementary** Figure 14).

Most loci did not have colocalized eQTLs, however, and we utilized chromatin loops in hepatocytes to annotate more distal target genes of MASLD loci. Overall, we linked 12 MASLD loci with caQTLs to putative target genes in hepatocytes using chromatin loops (**Figure 4E**), including those not previously prioritized as candidate genes (*44*). For example, at the 12q13 locus, the hepatocyte cCRE affected by MASLD variants interacted with the promoter region of *KRT8/KRT18,* which are keratins that form the intermediate filament cytoskeleton in hepatocytes (**Figure 4F**). Hepatocyte chromatin state at this locus further supports *KRT8/KRT18* as candidate genes, as other previous implicated genes such as *KRT78* are in broad regions of repression (**Figure 4F**). At the 8p23 locus, the hepatocyte cCRE affected by MASLD variants interacted with the *PPP1R3B* promoter over 200kb distal (**Figure 4G**). Additional interactions between associated cCREs and distal target genes included the 19q13 locus where cCREs interacted with the *CEBPA* promoter (**Supplementary** Figure 15), and the 22q12 locus where cCREs interacted with the *XBP1* promoter.

Finally, we prioritized candidate causal variants underlying genetic effects on hepatocyte cCRE activity at MASLD loci. We created sequence-based models of hepatocyte chromatin accessibility using chromBPNet (*53*), and then used models to predict the functional effects variants at each locus with a caQTL. At almost all (24/26) MASLD loci with hepatocyte caQTLs, at least one fine-mapped variant was predicted to have functional effects on hepatocyte cCRE activity (**Figure 4E, Supplementary Table 14**). For example, at the 12q13 locus, the most likely causal variant rs9668670 (PPA=.60) was predicted to disrupt hepatocyte cCRE activity, where the MASLD risk allele of this variant increased hepatocyte cCRE activity and had higher predicted accessibility (**Figure 4F**). At the 8p23 locus, likely causal variant rs4841132 (PPA=.99) was predicted to disrupt hepatocyte cCRE activity where the MASLD risk allele decreased cCRE activity and had lower predicted accessibility (**Figure 4G**). At both loci, candidate variants mapped within sequence motifs for HNF1A, a key regulator of hepatocytes, among other TFs (**Figure 4F,G**).

Overall, these results provide detailed annotation of MASLD loci directly affecting hepatocyte regulation and candidate functional variants and target genes of variant activity at these loci.

### Changes in hepatocyte sub-clusters in progression to MASH

We finally sought to understand cellular heterogeneity within hepatocytes and the role of hepatocyte heterogeneity in progression to MASH. We identified five distinct sub-clusters of hepatocytes, which we annotated based on multiple lines of evidence including gene expression profiles, spatial localization, and abundance in MASLD (**Figure 5A-D)**. This included three hepatocyte sub-clusters based on zonation within the liver including Zone 1 (Peri-portal), Zone 2 (Mid-lobular), and Zone 3 (Central-lobular) cells. For example, Zone 1 hepatocytes had increased expression of marker genes (*G6PC, GLS2, PCK1*) and pathways related to insulin signaling and localized around the portal vein, while Zone 3 hepatocytes had increased expression of known markers (*CYP2E1, ADH4*) and lipid metabolism genes and pathways and localized around the central vein (**Figure 5B,C**). Hepatocyte zonation was preserved in MASL and Fib- MASH hepatocytes, like that observed in normal livers, although there were increased Zone 1 and Zone 3 hepatocytes in MASL (**Figure 5D**). We also observed altered abundance of hepatocyte zones in MASH livers, most notably a decrease in Zone 1 and 3 hepatocytes in Fib+ MASH and higher fibrosis score (**Figure 5C,D**).

**Figure 5.**
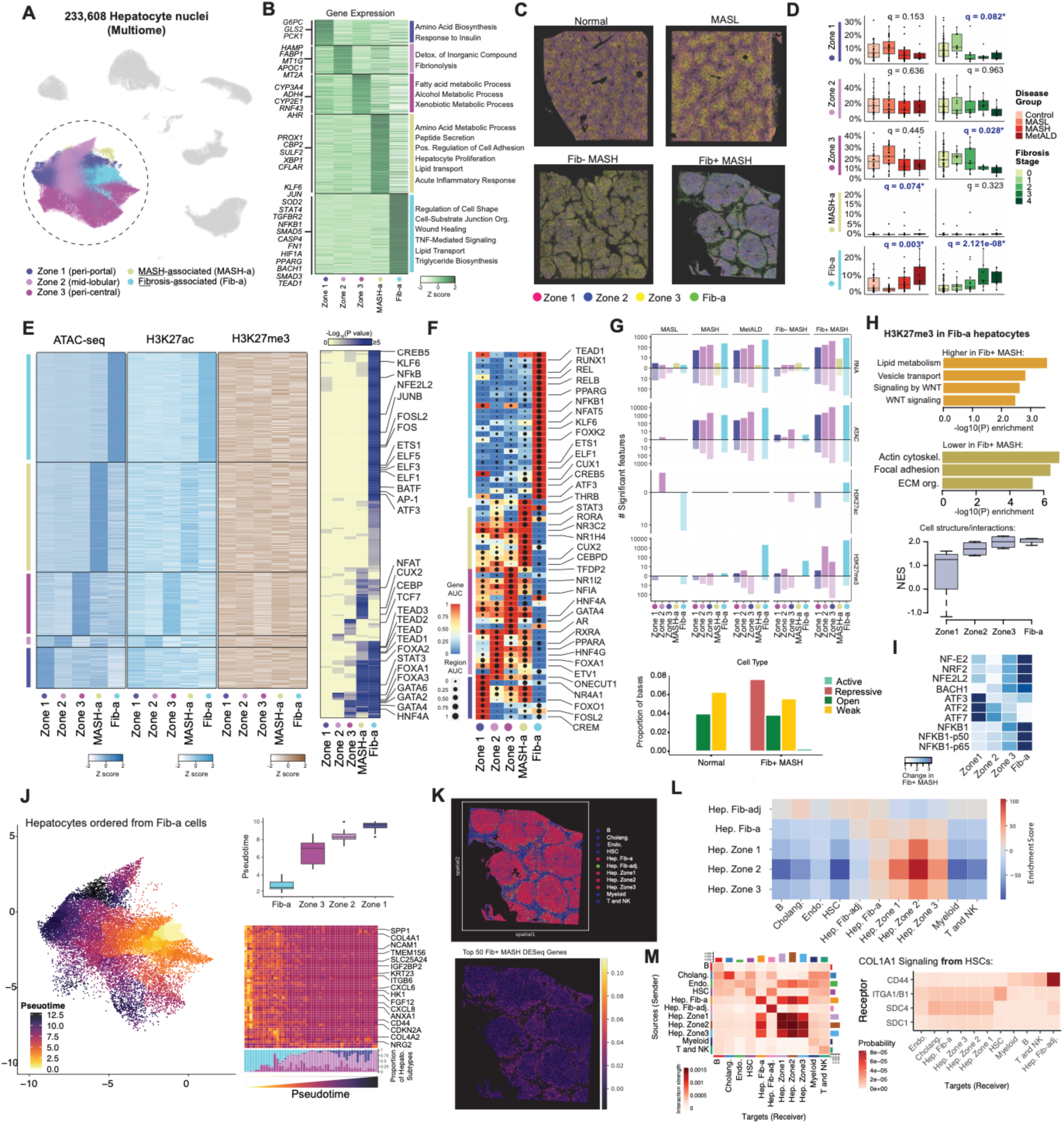
Heterogeneity in hepatocytes and changes in MASLD. (A) Subclusters of hepatocytes identified in single cell multiome and Paired-Tag data including zone 1 (peri-portal), zone 2 (mid-lobular), zone 3 (peri-central) populations, a MASH-associated (MASH-a) population, and a Fibrosis-associated (Fib-a) population. (B) Genes with specific expression in each hepatocyte sub-cluster (left) and biological pathways enriched in subcluster-specific genes (right). (C) Annotation of hepatocyte sub-clusters in spatial transcriptome profiles from representative examples of normal, MASL, Fib- MASH and Fib+ MASH livers. (D) Proportions of cells in hepatocyte subclusters across different MASLD states and fibrosis score. (E) Hepatocyte cCREs with activity specific to each sub-cluster in accessible chromatin, H3K27ac and H3K27me3 profiles (left) and TF sequence motifs enriched in subcluster-specific cCREs (right). (F) TF GRNs with increased activity in each hepatocyte sub-cluster. (G) Features for each modality with changes in activity in MASLD states within each hepatocyte sub-cluster (top) and the proportion of bases within H3K27me3 bins altered in injured hepatocytes in Fib+ MASH in each chromatin state (bottom). (H) Biological pathways enriched in H3K27me3 bins with increased and decreased activity in Fib-a hepatocytes in Fib+ MASH (top), and enrichment of cell structure/interaction pathways in Fib+ MASH in each hepatocyte sub-cluster (bottom). (I) Sequence motifs enriched in cCREs with increased activity in Fib+ MASH in each hepatocyte sub-cluster. (J) Pseudo-time ordering of hepatocytes from the Fib-a sub-cluster (left), where Zone 3 hepatocytes are ordered closest to Fib-a cells compared to other hepatocyte zones (top right). Expression of the top genes up-regulated in Fib+ MASH in hepatocytes and proportion of each hepatocyte subcluster across pseudo time bins (bottom right). (K) Cells in spatial transcriptomics of a Fib+ MASH donor colored by the joint expression of the top genes up-regulated in Fib+ MASH. (L) Cellular neighborhood analysis showing enrichment of each liver cell type and hepatocyte sub-cluster for proximity to other cell types. (M) Strength of overall interactions between pairs of liver cell types and hepatocyte sub-clusters based on predicted cell-cell signaling in Fib+ MASH (left), strength of interaction between ligand expressed in HSCs (*COL1A1*) and specific receptors expressed in liver cell types and sub-clusters in Fib+ MASH (right).

Outside of zonation-based heterogeneity we observed two additional sub-clusters of hepatocytes. The first sub-cluster preferentially expressed genes implicated in hepatocyte proliferation, identity and function such as *PROX1, XBP1, CBP2,* as well as pathways related to proliferation, metabolism, inflammation and lipid transport. This sub-cluster had generally low abundance (mean=1.2%), with several exceptions, and had significantly increased proportion in MASH compared to normal livers, and therefore we termed this sub-type ‘MASH-associated’ (MASH-a) hepatocytes (**Figure 5A-D, Supplementary Table 3**). The second sub-cluster preferentially

expressed many inflammatory and stress-related genes (*NFKB1, STAT4, HIF1A, SOD2, TEAD1, TGFBR2*) and inflammation, cell organization and structure, and lipid transport pathways (**Figure 5B**). This sub-cluster was present in all livers (mean=11.1%) and had significantly increased proportion in Fib+ MASH and severe fibrosis compared to normal livers (**Figure 5D, Supplementary Table 3**). We also observed localization of this sub-cluster near areas of liver fibrosis in MASH compared to normal and MASL livers (**Figure 5C**). Therefore, we termed this sub-type ‘fibrosis-associated’ (Fib-a) hepatocytes There was an inverse correlation between Fib- a hepatocyte proportion and Zone 3 (rho=-.47, p=7.2x10^-6^), and to a lesser degree Zone 1 (rho=- .32, p=.0022) across livers, with no relationship with Zone 2 hepatocytes (rho=.05).

Next, we determined the transcriptional regulatory programs driving hepatocyte sub-cluster identity. We identified 1,010 hepatocyte cCREs with accessible chromatin profiles highly specific to each hepatocyte sub-cluster, and these cCREs had concordant specificity in H3K27ac activity (**Figure 5E, Supplementary Table 15**). Interestingly, we also observed a decrease in repressive H3K27me3 activity in Zone 3 cells among the cCREs with Zone 3-specific activity. Zone-specific cREs in hepatocytes were enriched for specific TF motifs such as GATA and TCF7 motifs in Zone 3 hepatocytes, and FOXA motifs in Zone 2 hepatocytes. The MASH-a and Fib-a specific cCREs were also enriched for TF sequence motifs, where Fib-a hepatocytes showed enrichment for many motifs including FOS/JUN, NFKB, TEAD, ELF, ETS1, and ATF family TFs and MASH-a hepatocytes were enriched for FOXA, CEBP, PPAR and steroid hormone (GR, AR) motifs **(Figure 5E, Supplementary Table 16**). We further identified TF GRNs with increased activity in each hepatocyte sub-cluster which implicated similar TFs driving zonation identity including HNF4A and GATA4 in Zone 3, FOXA1, PPARA and RXRA in Zone 2, and FOXO1 and NR4A1 in Zone 1 hepatocytes. Numerous TF GRNs were also highly active in Fib-a hepatocytes compared to other hepatocyte sub-types including NFKB1/REL, TEAD1, ETS1, ELK1, KLF6, CREB5, CUX1, and ATF3, among others (**Figure 5F**).

### Changes in hepatocyte heterogeneity in progression to MASH

We sought to define genomic changes within each hepatocyte sub-cluster in progression to MASH. Overall, there were dramatic changes in each sub-cluster in MASH/MetALD as well as in Fib+ MASH, and few changes in MASL and Fib- MASH, compared to normal livers (**Figure 5G, Supplementary Table 17, Supplementary Data**). Zone 3 and Fib-a hepatocytes showed the largest number of changes in gene expression and cCRE activity in Fib+ MASH compared to other sub-clusters (**Figure 5G**). Notably, we also observed a substantial number of changes in the repressive mark H3K27me3 across genomic regions (15kb bins) in Fib-a hepatocytes, suggesting broadly altered repression profiles within hepatocytes in Fib+ MASH. Defining the chromatin state of H3K27me3 bins altered in Fib+ MASH further confirmed an increase in repression in fibrosis-associated hepatocytes (**Figure 5G**). Specific biological pathways were enriched in genes overlapping regions with altered H3K27me3 activity in Fib-a hepatocytes, with increased repression at lipid metabolism, transport, and Wnt signaling pathways leading to decreased expression, and decreased repression at cell structure, adhesion, ECM interaction and cytokine signaling genes leading to their activation (**Figure 5H**). The Fib-a hepatocytes also had the largest increases in expression of genes within selected cell structure, adhesion and ECM pathways in Fib+ MASH relative to other sub-clusters (**Figure 5H**).

We next characterized changes in cCRE activity in MASH within each hepatocyte sub-cluster. We identified TF sequence motifs enriched in cCREs with up- and down-regulated activity in Fib+ MASH and identified motifs with differential enrichment across hepatocyte sub-clusters. For example, among cCREs up-regulated in Fib+ MASH, there was stronger enrichment of NFKB- related motifs as well as motif for oxidative stress-related TFs such as NFE2L2, NF-E2 and NRF2 in Fib-a and Zone 3 hepatocytes (**Figure 5I, Supplementary Table 18**). By comparison, there was stronger enrichment of ATF motifs as well as several FOS/JUN-related motifs in Zone 1 hepatocytes in Fib+ MASH (**Figure 5I, Supplementary Table 18**). We observed similar patterns among cCREs down-regulated in Fib+ MASH, with strong enrichment of SP and KLF TF family motifs in Fib-a and Zone 3 hepatocytes, and enrichment of STAT motifs in Fib-a hepatocytes, and few enriched motifs in other cell states.

The abundance of Fib-a hepatocytes inversely correlated with Zone 3 and Fib-a hepatocytes repressed Zone 3-related processes in MASH, and we hypothesized that Fib-a hepatocytes may preferentially arise from Zone 3 cells. We ordered hepatocytes in Fib+ MASH livers on a trajectory starting from Fib-a cells using pseudo-time analyses (**see Methods**), which revealed that Zone 3 cells were generally ordered more closely to Fib-a cells compared to Zone 1 and 2 cells (**Figure 5J**). We next ordered the expression of genes up-regulated in hepatocytes in Fib+ MASH, including cytokines (*CXCL8, CXCL6*), heat shock proteins (*HSPB8*), integrins (*ITGB6*), senescence-related genes (*CDKN2A*), and keratins (*KRT23*), along this trajectory. Overall, genes up-regulated in Fib+ MASH showed a gradient of expression that was highest in Fib-a and Zone 3 hepatocytes. Notably, these genes had particularly high expression within a subset of Fib-a hepatocytes. Furthermore, the hepatocytes expressing these genes most up-regulated in Fib+ MASH showed highly specific localization around the areas of fibrotic injury in Fib+ MASH livers (**Figure 5K**). We thus termed this subset of Fib-a hepatocytes adjacent to fibrotic injury in Fib+ MASH as ‘Fib-adj’ hepatocytes.

Finally, we hypothesized that changes in hepatocytes in MASH may be driven through altered signaling from cells in fibrotic areas in the liver. Based on neighborhood analysis, Fib-adj hepatocytes were specifically localized to cell types in fibrotic areas of the liver such as HSCs and immune cells in Fib+ MASH, while other hepatocyte sub-clusters mapped to different areas of the liver (**Figure 5L**). We performed cell-cell interaction analyses to identify signals altered between cell types in Fib+ MASH livers using CellChat (*54*, *55*). Overall, there was evidence for signaling from HSCs specifically to Fib-adj hepatocytes relative to other hepatocyte sub-clusters, while hepatocyte zones had increased signaling amongst each other as well as from endothelial cells (**Figure 5M**). The strongest signal from HSCs to Fib-adj hepatocytes involved collagen (*COL1A1*) and *CD44*, which is a cell surface adhesion molecule implicated in fibrogenic injury response and hepatocellular carcinoma (*56*, *57*) and is up-regulated in hepatocytes in MASH, while other hepatocytes had signaling to other receptors.

## Discussion

Single cell studies of normal and MASLD livers have largely consisted of gene expression profiling from relatively few donors (*16*, *29*, *58–63*). Combining multiple single cell epigenomic modalities, including accessible chromatin, active and repressive histone modifications, and 3D chromatin interactions, with single cell and spatial transcriptomics across many human livers revealed unprecedented insight into the regulatory programs of liver cell types as well as transcriptional drivers of progression to MASH. Compared to cCREs defined from accessible chromatin alone, incorporating histone modifications annotated chromatin state in liver cell types for a much larger proportion of the human genome. Furthermore, chromatin state and chromatin interactions provided functional context to cCRE activity and revealed distinct properties and disease risk enrichments of cCREs in different chromatin states within liver cell types. The additional epigenomic modalities also enabled an improved annotation of genetic loci associated with MASLD risk in liver cell types, including candidate variants not annotated in previous cCRE catalogs and distal target genes of risk variant activity. Collectively, multimodal profiling enabled a more comprehensive understanding of how liver cell types are regulated and the regulatory programs driving MASLD risk and pathogenesis.

Only ∼20% of patients with MASL ultimately develop MASH, and the factors driving progression from MASL to MASH are not well understood. Hepatocytes respond first to metabolic injury early in disease and are key triggers of changes in HSCs and other cell types that lead to MASH and fibrosis. Our results revealed significant, stepwise differences in the genomic profiles of hepatocytes in MASL, low fibrosis MASH, and high fibrosis MASH, and uncovered key transcriptional drivers of these distinct profiles. Hepatocytes in MASL were marked by increased lipid metabolism, synthesis and uptake as well as increased activity of TF GRNs regulating these processes such as HNF4A, RORA and CUX2. In low fibrosis MASH, lipid-related pathways and GRNs remained active, while new programs associated with epithelial remodeling and adhesion regulated by TFs such as TEAD1, KLF6, ATF3, CREB5, and RELB were also activated. Finally, the transition to high fibrosis MASH resulted in a reduction in lipid metabolism and associated TF GRNs with a marked increase in inflammatory and fibrotic pathways regulated by TFs such as NFKB and NFAT5. Among these changes, lipid-related networks increased in MASL and Fib- MASH were most enriched for MASLD-associated variants, therefore linking genetic risk to processes up-regulated in the early stages of disease. We speculate that a more beneficial hepatocyte response to lipids may prevent further exacerbation of disease to fibrosis, which may be leveraged to predict progressors, identify biomarkers and inform therapeutic discovery.

Severe fibrosis in the liver was also marked by dramatic transitions in cellular sub-types of hepatocytes, including a loss of hepatocyte zonation and an increase in specific fibrosis-associated cells. Cellular heterogeneity in hepatocytes in MALSD has been previously reported, although largely at the transcriptional level (*16*, *23*, *63–68*). Our results revealed transcription factors and regulatory networks underlying zonation-based identity and other cellular heterogeneity in hepatocytes as well as transcriptional drivers of changes within hepatocyte zones and sub-types in MASH. Notably, the fibrosis-associated hepatocyte population had dramatically altered repressive domains marked by H3K27me3, including increased repression at lipid metabolism genes and loss of repression at epithelial remodeling and fibrosis-related genes. As this population was most closely related to Zone 3 hepatocytes, which themselves exhibited large genomic changes in MASH, we speculate that epigenetic remodeling drives a transcriptional switch from Zone 3 cells toward the fibrosis-associated state. The strong impact of fibrosis on transcriptional and epigenetic changes in hepatocytes was further illustrated by the localization of a specific subset of hepatocytes with dramatic changes in severe fibrosis directly at the sites of fibrosis. This hepatocyte subset also exhibited distinct cell-cell interactions with HSCs, notably involving *CD44* which is also implicated in hepatocellular carcinoma (*69*), and downstream signaling from these interactions may help drive transition to the fibrosis-associated state.

Combining cell type-specific gene regulatory programs in the liver with genetic association data revealed mechanisms of individual loci associated with MASLD. At a large fraction of MASLD loci, we detailed the regulatory logic of risk variant activity in hepatocytes, implicating specific variants, cCREs, transcriptional regulators, and genes as causally involved in disease. Through both QTL mapping and sequence-based models, we also uncovered the direction of effect of variants on hepatocyte regulatory activity, which may help inform strategies to modify element or gene activity to protect hepatocytes during disease. Chromatin loops highlighted candidate genes in hepatocytes not prioritized in a previous study (*44*), including those distal to risk variants, and revealed novel targets for MASLD. Candidate genes included *PPP1R3B,* where knockout in both mice and humans is associated with increased steatosis and hepatic fat (*70*), and *CEBPA,* a key hepatic TF involved in glucose and lipid metabolism implicated in liver fibrosis (*71*), as well as other genes such as *KRT8/18* and *XBP1* involved in hepatocyte function and

In summary, our multimodal study provided the most comprehensive annotation to date of gene regulatory programs of liver cell types and cellular heterogeneity which enabled new mechanistic insight into drivers of MASLD progression and uncovered novel targets for therapeutic discovery.

## Acknowledgements

The authors would like to thank Lifesharing OPO. We thank the members of the Gaulton, Ren, Brenner, and Kisseleva labs for comments and feedback, and the University of California Institute for Genomic Medicine (IGM).

## Author contributions

W.E. performed single cell and genetic analyses and wrote the manuscript. L.C. performed Paired-Tag and droplet Hi-C data generation and wrote the manuscript. Y.X. performed single cell analyses and wrote the manuscript. Q.Y. performed sample preparation and single cell experiments. C.M. performed single cell and spatial transcriptomics analyses. H.M., R.L., S.C., M.K., L.T., M.T, K.D., R.M. performed single cell analyses. E.F. and N.C. contributed to single cell analyses. S.S. and H.Y.K. processed and annotated human liver samples and interpreted results. J.L. generated spatial transcriptomics data. S.M. performed analyses to create sequence models of chromatin. T.V. and J.O.B. performed spatial analyses. D.L., C.S. and T.W. developed the single cell browser. M.V. contributed to droplet Hi-C analysis J.D., E.E., J.B., and M.M performed sample preparation and single cell experiments. A.L. and T.L. performed single cell data generation. A.D. supervised single cell and spatial data generation. A.W., D.B., T.K., B.R., K.J.G. supervised the study and revised the manuscript. K.J.G., B.R., T.K. and D.B. conceived the study, obtained funding, and wrote the manuscript.

## Data and materials availability

The raw data generated in this project will be made available in GEO upon publication. Processed data are available at https://epigenome.wustl.edu/MASLD/. Supplementary data is available in Zenodo doi:10.5281/zenodo.15298484. Code for analyses performed in the project is available at https://github.com/Gaulton-Lab/FNIH.Liver.

## Supplementary Materials

Materials and Methods Supplementary Figures 1-15

Supplementary Tables 1-18 Supplementary Data References

## Materials and methods

### Donor collection and information

Deidentified livers (IRB 171883XX, certified by HRPP Director and IRB Chair as not human subjects according to the Code of Federal Regulations, Title 45, part 46 and UCSD Standard Operating Policies and Procedures) from 87 donors were obtained via Lifesharing OPO, which provided the informed consent, laboratory tests (ALT, AST, biochemistry, cell counts, liver biopsy, serology, and others), as well as patient history (cause of death, age, BMI, gender, and underlying diseases, history of alcohol consumption). Livers were graded by a pathologist using a double-blinded method and identified as normal, MASL, and MASH. Based on the donor history, biochemical, and histological evaluation, livers were graded as normal, MASL or MASH. MASH Clinical Research Network (MASH CRN, 2005) methodology was used for evaluation of livers including the degree of steatosis, steatohepatitis, inflammation, and fibrosis. A NASC/CRN score of 5 was used as the cut off for diagnosis of MASH. Livers with scores of <3 were diagnosed as not having MASH. Representative images of normal, MASL, and MASH livers are shown in **Supplementary** Figure 4.

### Single cell multiome assays

Snap frozen liver samples (5x5 cm^2^) from each donor were used for single cell assays. For each pool, we selected between 15 to 30 donors to represent all MASLD groups and sexes. For each donor, approximately 20-30 mg of snap frozen tissue (tissue samples measuring approximately 3 mm X 3 mm X 3mm) was sectioned on dry ice using pre-chilled scalpels and collected in a pre-chilled 5ml LoBind tube on dry ice. Sectioned tissue pools were stored at -80°C freezer. The nuclei suspension was then filtered through 100-μm and 30-μm CellTrics™ Filters and centrifuged for 5 min at 500g at 4 C. Anti-Nucleus MicroBeads (Miltenyi Biotec) were used for debris removal and enrichment of nuclei. The nuclei suspension was centrifuged for 5 min at 500g at 4 C. After resuspension in sort buffer (1X PBS, 1X protease inhibitor, 1.5 U μl− RNasin® Ribonuclease Inhibitor and 1% BSA), nuclei were stained with 2 μM 7-AAD in sort buffer for 10 min on ice and were sorted by fluorescence-activated nuclei sorting with an SH800 cell sorter for single nuclei. Nuclei were collected in collection buffer (1X PBS, 5X protease inhibitor, 5 U μl^−1^ RNaseOUT, 5 U μl^−1^ SUPERaseIn inhibitor, and 5% BSA) and centrifuged for 10 min at 500g at 4C. The nuclei pellet was resuspended in permeabilization buffer (1mM DTT, 0.2% IGEPAL-CA630, 1X cOmplete EDTA-free protease inhibitor, 1.5 U μl−1 RNasin and 5% BSA in PBS), incubated on ice for 2 min and centrifuged for 5 min at 500g and 4°C and resuspended in 1x nuclei buffer. Suspension of 30,000 permeabilized nuclei were each loaded onto 8 lanes and processed using of 10X Genomics Controller following manufacturers recommendations. 10x Genomics Epi Multiome ATAC + Gene Expression assays were processed following manufacturers recommendations. Generated libraries were sequenced at the Institute for Genomic Medicine at UC San Diego using NovaSeq X Plus (Illumina) sequencer.

### Single cell droplet paired-Tag assays

To obtain joint profiles of histone modifications and transcriptomes from single cells in liver samples, Droplet Paired-Tag was performed as previously described, with minor modifications. For this study, we performed separate assays for antibodies targeting the histone modification marks H3K27ac (Abcam, ab4729, polyclonal) and H3K27me3 (Abcam, ab192985, recombinant). Briefly, liver tissues from 16–24 different donors were pooled in equal weights, and nuclei were extracted following the same protocol used for our 10x Genomics single-cell multiome data generation. The nuclei were stained with 0.2mg/ml Hoescht and were sorted via fluorescence-activated nuclei sorting using an SH800 cell sorter (Sony) to isolate single nuclei. The isolated nuclei were collected and counted using a cell counter (RWD C100-Pro) with DAPI staining. We aliquoted 500,000 nuclei to initiate reactions in individual tubes for each histone modification mark and replicate. Two replicates were included for each tissue pool and histone modification.

First, nuclei were permeabilized with OMNI buffer and incubated with 2 μL PA-Tn5 (0.4 mg/mL) and 2 μg antibody in MED#1 buffer overnight at 4 °C. PA-Tn5 and antibodies were then removed by washing with MED#2 buffer. Tagmentation was carried out in MED#2 buffer supplemented with 10 mM MgCl2 (Invitrogen, AM9530G) at 550 r.p.m. and 37 °C for 60 minutes in a ThermoMixer (Eppendorf), and the reaction was terminated by adding 2× stop solution. The nuclei were washed in 1× nuclei buffer and counted again. From each tube, 25,000–40,000 nuclei were aliquoted and used for a single droplet generation reaction with the Chromium Next GEM Single Cell Multiome kit (10x Genomics, 1000283). Finally, DNA and RNA library amplification was performed according to the Chromium Single Cell ATAC Library kit manual, with the following modifications: the starting amount of preamplification product for DNA library amplification was doubled, 13 amplification cycles were used for the DNA library, and a different SPRI bead size selection were applied for the DNA library (50 μL + 105 μL). The libraries were sequenced at the UCSD IGM using the Illumina NovaSeq X.

### Single cell droplet Hi-C assays

To obtain cell-type specific chromatin architecture, we performed Droplet Hi-C on all the livers from different donors, as previously described. Liver tissue from 16–24 donors were pooled together, and nuclei were extracted within the same reaction. Of the 87 donors, one donor did not have enough tissue available for pooling and was excluded from Droplet Hi-C pools. Nuclei were fixed with formaldehyde (Sigma, 47608-250ML-F) at a final concentration of 1% or paraformaldehyde (Electron Microscopy Sciences, 15714) at a final concentration of 2%, and then quenched with glycine solution at a final concentration of 0.2 M. Nuclei were permeabilized with lysis buffer on ice. Then, the nuclei were treated with 0.5% SDS (Promega, V6551), incubated at 62 °C for 10 minutes on a ThermoMixer, and then quenched with Triton X-100 (Sigma, 93443). Chromatin in the nuclei was sheared using three restriction enzymes: DpnII (NEB, R0543L), MboI (NEB, R0147M), and NlaIII (NEB, R0125L), at 37 °C for 90 minutes on a ThermoMixer at 550 r.p.m. The three enzymes were then deactivated at 65 °C for 20 minutes, and the mixture was cooled to room temperature. The sheared chromatin was further ligated using T4 DNA ligase (NEB, M0202L). All enzymes were removed from the nuclei, which were then suspended in 1% BSA in 1xPBS with 7AAD (Invitrogen, A1310) and incubated on ice for 1 hour.

The nuclei were washed three times with 1× nuclei buffer. Approximately 20,000 nuclei were aliquoted into each PCR tube, and one reaction was performed using the Chromium Next GEM Single Cell ATAC Reagent Kits v2 (10x Genomics). The tagmentation incubation time was 60 minutes, and the index PCR elongation time was extended from 20 seconds to 1 minute. Double-sided size selection was adjusted to 1.14× SPRIselect to remove only small fragments. For each liver sample pool, we performed at least two replicates, with each replicate involving 1–3 10x Genomics reactions. The libraries were sequenced at UCSD IGM using the Illumina NovaSeq X.

### Genotyping arrays and imputation

From all donors with liver single cell profiles, we sectioned 10-15mg of frozen tissue and extracted genomic DNA from each donor using the Monarch Genomic DNA Purification Kit (NEB #T3010L). Samples were eluted in 70-100uL UltraPure Distilled Water (Invitrogen 10977-015), and speed vacuumed to a concentration of 50ng/uL when necessary. Samples were genotyped using the Illumina InfiniumCoreExome-24v1-4_A1 array at the UCSD IGM Genomics Center.

Cluster positions and SNP statistics were extracted from the array data using GenomeStudio v2.0.5. Genotypes were converted to plink format using GenomeStudio v2.0.5 with the plink input report plugin using an hg38 cluster file. Samples from different arrays were merged using plink v1.9, variants were filtered based on a minimum missing call rate of 0.05, minor allele frequency of at least .01, and Hardy-Weinberg equilibrium p-value greater than 1e-5 before generating VCF and frequency files. We then used the HRC-1000G-check-bim.pl script from the McCarthy Group Tools (https://www.chg.ox.ac.uk/~wrayner/tools/, v4.3.0) to filter and prepare variants for imputation.

Genotypes were imputed on using TOPMed r3 panel using the TOPMed Imputation Server(*1*). Imputation was performed using the Array Build Hg38 with r^2^ filter off and phasing using Eagle (v2.4). Following imputation, variants were filtered by imputation quality (r^2^>0.9 and minor allele frequency (MAF) > 0.01.

### Spatial transcriptomics (Visium HD) assays

To minimize potential RNAse contamination dissection surfaces were cleaned with 70% EtoH sterile water was used for floating sections and clean new blades were use for microtome sectioning. FFPE blocks were hydrated for 5-10min on top of ice chilled flat metal surface and sections were cut at 5um thickness mounted to a Superfrost™ Slides (Fisher Scientific). Slides were dried for 2 hours in the oven at 42C after sectioning and stored at room temperature with desiccators. All sections were used for experiments within 2 weeks of mounting. To assess the RNA integrity of the tissue, we assayed DV200 fragments. Slices of 3-5 pieces at 10um thick ribbon slices were saved prior to mounting. RNA was isolated as recommended in PureLink™ FFPE Kit (Invitrogen). All samples with DV200 fragments greater than 60% are used for the Visium HD experiments.

Deparaffinization and H&E staining conditions were carried out as recommended by 10X Genomics Visium HDFFPETissuePrepHandbook_RevA CG000684. Briefly, H&E images were acquired by Olympus VS200 Slide Scanner at 20X magnification. Immediately after imaging, the slides are decrosslink and hybridize overnight with Visium Human Transcriptome Probes v2 >18,000 genes, >54,000 probe pairs. After permeabilizing and barcoding, the hybridized RNA was transferred to a Visium Slide slide 6.5mmX6.5mm oligo capture area. The ligated and barcoded transcripts were then subjected to amplification and indexing for library construction. Paired-end, dual indexed sequencing conditions were Read1: 43 cycles, i7 Index: 10 cycles, i5 Index: 10 cycles and Read 2S: 50 cycles. The libraries were sequenced at the UCSD IGM using Illumina NovaSeq X.

### Single cell multiome clustering and annotation

The 10x Multiome fastqs were aligned and processed via Cell Ranger Arc v2.0.2. For each library we generated a ATAC count matrix of 5kb windows tiling the human genome by removing duplicate reads from the atac_possorted_bam.bam file from Cell Ranger, converted to tagAlign format, and quantified counts with bedtools v2.27.1. For each library, sample pools were deconvoluted using demuxlet(*2*) (v2) (https://github.com/statgen/popscle) using a VCF subset to reads in open chromatin sites(*3*) and samples in that pool, a merged and sorted bam made from the Cell Ranger gex_possorted_bam.bam and atac_possorted_bam.bam, and the list of barcodes used to make the 5kb window matrix.

The raw RNA count matrix was loaded into Seurat v5.0.3, subset to the list of barcodes used to make the 5kb windows matrix, and SoupX v1.6.2 (*4*) was used to remove ambient RNA. The 5kb windows matrix was added to the object as a ChromatinAssay using Signac v1.12.0 (*5*) and donor assignments were added based on the demuxlet outputs, filtering out any barcode called as a doublet. The resulting object was clustered using the weighted nearest neighbor (WNN) approach to check library quality.

Prior to merging libraries, the ATAC assay was subset to the 50k most variable windows. Cells were further filtered after merging all libraries together for <2% mitochondrial gene and TSS enrichment >2 as calculated by Signac(*5*). After clustering, we identified residual populations that appeared to be doublets based on high expression of markers from two different cell types and higher gene expression. To remove these potential doublets, we ran Amulet v1.1 (*6*) and re-clustered the data at high resolution and removed clusters with greater than 5% cells marked as doublets by Amulet.

The resulting object was clustered using the RNA modality and cell types were annotated based on the expression of canonical liver marker genes, such as Albumin (*ALB*) for Hepatocytes and collagen genes in stellate cells. A full list of genes used to annotate cell types is provided in **Supplemental Table 2.** One donor (HL180069) in the original sample pools, was excluded from downstream analyses due to concerns with sample quality based on consistently being an outlier in count data.

Peaks were called by cell type using MACS2 v2.2.7.1 (*7*) through Signac. An initial peak set across cell types was generated by iteratively clustering overlapping peaks, selecting the peak with the highest pileup score, removing other peaks directly overlapping that peak, and repeating until no overlaps remain. This consensus peak set was used to generate a new matrix using the FeatureMatrix command in Signac. The resulting data was re-clustered and final cell type annotations were identified based on expression patterns of canonical liver genes and marker genes in Seurat. A final set of consensus peaks were identified for cell types and sub-types using the following pipeline. ATAC-seq bam files were converted to tagAlign format and merged, then split by cell type or subtype using a barcode list. Peaks were called on the resulting split tagAlign file and a consensus peak set was generated across cell types and subtypes using the same iterative clustering approach described above. Finally, a barcode level count matrix was generated using FeatureMatrix and added back into the object and used for analyses.

### Preprocessing of Droplet Paired-Tag data

Fastq files from Droplet Paired-Tag experiments were demultiplexed using the mkfastq command in cellranger-arc v2.0.0. After demultiplexing, the histone modification and transcriptome modalities were aligned separately to the human reference genome (hg38) using the count command in cellranger-atac v2.0.0 for histone modifications or cellranger v6.1.2 for transcriptome. To ensure the inclusion of high-quality nuclei, we implemented a four-step filtering strategy: i) Histone Modification Data Filtering: At the sample level, histone modification data were aggregated, and peak calling was performed using MACS2 v2.1.2 (*7*) to identify narrow peaks (for H3K27ac) and broad peaks (for H3K27me3). High-quality barcodes were selected based on the number of fragments per cell and the Fraction of Reads in Peaks (FRiP) score. ii) Transcriptome Data Filtering: For RNA data, barcodes were filtered based on the number of unique molecular identifiers (UMIs) per cell. iii) Nuclei Matching: Pass-filter barcodes from both modalities were paired to select high-quality nuclei. iv) Donor Assignment: We used the genotype information from each donor to distinguish the barcodes associated with single or multiple donors as described above for multiome data. High-quality nuclei confidently assigned to a single donor were retained for downstream analyses. Finally, to address ambient RNA contamination, we used SoupX (*4*) to estimate and remove contamination for each library independently prior to clustering.

### Reference mapping of Droplet Paired-Tag data with 10X Multiome

We used the RNA modality from 10X Multiome assays as reference to annotate cell types from Droplet Paired-Tag data using Seurat v5.1.0 (*8*). In brief, Droplet Paired-Tag datasets were first normalized and scaled using SCTransform (*9*). Integration anchor cells was identified with the ‘FindTransferAnchors’ function to project the reference (10X Multiome) PCA structure to query (Droplet Paired-Tag) data. The first 30 principal components were then used for UMAP visualization and cell identity annotation on major cell type or cell subtype level using ‘MapQuery’ function with default parameters. For downstream analysis at the major cell type and cell subtype level, cells with a prediction score <0.8 were removed.

### Visualization of ATAC and histone modification tracks

Cell type- or condition-specific alignments for histone modification data were extracted from cellranger bam files using custom scripts. BigWig files were generated using the ’bamCoverage’ function in deepTools v3.5.1 applying the RPGC normalization method (*10*). Chromosome Y (chrY) was excluded from the normalization process.

For chromatin accessibility data, the shifted and extended tagAlign files originally used for peaks calling were converted back to bam files for BigWig generation. The bamCoverage function was used to generate BigWig files, employing the same normalization method RPGC and exclusion of chrY as applied for histone modification data.

### Preprocessing of Droplet Hi-C data

Droplet Hi-C data were processed following previously described protocols. Fastq files were demultiplexed using the mkfastq command in cellranger-atac v2.0.0. After demultiplexing, cellular barcode sequences were extracted and aligned to the whitelist using Bowtie v1.3.0 (*11*), and appended to the beginning of each read name to record cellular identity. Sequencing adapters were trimmed using Trim-Galore v0.6.10 (*12*), and reads were aligned to the human reference genome (hg38) with BWA-MEM v0.7.17 using the arguments ‘-SP5M’(*13*). Contact pairs were then parsed, sorted, and deduplicated using Pairtools v0.3.0 (*14*), with barcode information stored in separate columns within the pairs file.

High-quality nuclei in each library were selected based on the number of unique contact pairs per cell. Genotype information was then used for donor phasing, and nuclei confidently assigned to a single donor were retained. These nuclei were split into individual pairs files, each containing the complete contact information for a single nucleus. Imputation was conducted using scHiCluster (v1.3.5) at three resolutions: 100 kb for compartment analysis, 25 kb for domain analysis, and 10 kb for cell embedding and loop analysis (*15*).

### Reference mapping of Droplet Hi-C data

We applied a previously described method to reference-map and annotate Droplet Hi-C profiles using gene expression data generated from the same tissue and samples (*16*). Briefly, the single-cell gene associating domain (scGAD) score *Rij* was calculated as the total contact number across the gene body for gene *i* in cell *j* at 10 kb resolution. The chromatin interaction profiles were represented as a cell-by-gene scGAD score matrix, which has been shown to correlate with gene activity. This matrix served as the input for reference mapping using the snRNA-seq data.

For reference mapping, the RNA modality from 10X Multiome was used. The PCA model derived from the reference dataset was then applied to transform the scGAD score matrix, with features normalized and scaled to match the reference. Integration of the scGAD score matrix with the reference dataset was performed using canonical correlation analysis (CCA), and the integrated embedding was transformed and visualized using the pre-calculated UMAP embedding model from the reference dataset with Seurat function ‘ProjectUMAP’.

To annotate cell identities based on scGAD scores, we identified the 15 nearest neighbors in the reference RNA data for each Droplet Hi-C cell using PyNNDescent v0.5.6 with Euclidean distance as the metric (*17*). Distances were scaled and standardized, with the nearest neighbor receiving the highest score. The cell type identity of each Droplet Hi-C cell was inferred by assigning the cell type label that received the highest standardized score among its nearest neighbors.

### Identifying chromatin loops in liver cell types

We used scHiCluster to identify loops at 10kb resolution from imputed contact maps in selected major cell types. scHiCluster implements a modified version of SnapHi-C and performs loops calling between genomic distances of 50kb and 5Mb. In addition, we identified loop summits which represent loop anchors with the highest normalized signal against the global background within a 20kb window.

### Genome annotation using ChromHMM

We used ChromHMM to summarize all epigenetic modalities (ATAC, H3K27ac, H3K27me3) and annotate the genome (*18*). To generate ChromHMM model, we first split the fragments files from cellranger-atac output by cell type. The resulting bed files were binarized using the binarizeBed function of ChromHMM, with the ‘-center’ flag specified. A model with states number set to 5 were

learned using the LearnModel function with default parameters. Chromatin states were manually annotated based on the likelihood of epigenetic mark associated with each state. It should be noted that the states identified here may differ from those in other studies such as the Epigenome Roadmap project due to the more limited number of histone marks available.

### Annotation of cCREs using ChromHMM

To annotate candidate cis-regulatory elements (cCREs), we determined the overlap between ChromHMM segments and the identified cCREs from 10X Multiome data using “bedtools intersect” command (*19*). A ChromHMM state was assigned to a cCRE if it overlapped with at least 50% of the cCRE region. In rare cases (<1%) where a cCRE exhibited equal proportions of multiple chromatin states, chromatin state was randomly assigned. Since ChromHMM states are called using 200 bp bins while the median length of cCREs is 300 bp, the inclusion of more than two chromatin states for a single cCRE is very rare and was not considered in this study.

### Changes in sample cell type proportions

To compare the proportion of cell types and cell sub-types across donors with different disease stages, we first extracted and square root scaled the proportion of each cell type per donor. We arranged disease classifications along a continuous scale with control as 0, MASL as 1, MASH as 2 and MetALD as 3. We also considered fibrosis stage and steatosis grade. Then, we trained generalized linear models to predict the scaled proportion of each cell type using condition, fibrosis stage, or steatosis, along with scaled donor age and BMI. We extracted the coefficient and p-value associated with each covariate from the trained model and performed Benjamini Hochberg multiple test correction to identify donor covariates significantly predictive of cell type proportions. Additionally, we performed an ANOVA on the donor condition information to identify differences in cell type proportions between disease groups. Two donors (HL170065, HL201024) were excluded from these analyses because they had outlying values for BMI, over 3 standard deviations from the mean of all donors. When comparing the proportions of hepatocyte sub-types across donors, we calculated the proportion of each sub-type relative to the total number of hepatocytes.

### Defining gene regulatory programs in each cell type

Cell type marker genes were identified by generating cell type pseudobulk profiles for each donor, calculating the average transcripts-per-million (TPM) for each gene across donors, and calculating entropy using the Entropy function from the DescTools package in R. We then considered markers of a cell type as those with the lowest entropy values and at least TPM>1 in the cell type.

### Defining cell type specific cCREs

To identify cell type- or subtype-specific cCREs, we first performed a regression-based differential accessibility test for each cluster using SnapATAC2 v2.7.0 (*20*). Differential cCREs were then assigned to specific groups using a described method based on a specificity score derived from Jensen-Shannon divergence (JSD) (*21*). Briefly, we calculated and normalized the fraction of accessibility for each differential cCRE across clusters to account for differences in complexity between groups. The normalized accessibility values served as input for JSD calculation. JSD was computed between the accessibility distribution of each cCRE and its reference distribution using the R package “philentropy”. The reference distribution (*d*) for a given cCRE in given group (*g1*) is defined as:

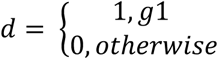

The Jensen Shannon-based specificity score (JSS) was defined as 1 − *JSD*. We calculated JSS for every cCRE for each group. We consider JSS from cCREs that are not identified as differential from likelihood ratio test as a background distribution, and JSS from differential cCREs with positive values to be true positives. We set an empirical false discovery rate (FDR) cutoff at where type I error no more than 5%. We defined those cCREs that can be assigned to only one cell type or subtype as cell type- or subtype- specific cCREs. Finally, cCREs uniquely assigned to a single cell type or subtype based on their JSS were defined as cell type- or subtype-specific cCREs.

### Identification of cis-regulatory modules

We employed non-negative matrix factorization (NMF) to classify cCREs into modules based on their relative accessibility across cell types (*22*). Using the NMF implementation in scikit-learn (*23*), we decomposed the cell-by-cCRE matrix *V* (*N×M*, where *N* = cCREs, *M* = cells) into a coefficient matrix *H* (*R×M*, where *R* = number of modules) and a basis matrix *W* (*N×R*). The basis matrix represents module-associated cCREs, while the coefficient matrix *H* describes the cell type components and their weights within each module. To determine an appropriate rank *R*, we evaluated multiple metrics including ‘Sparseness’ and ‘Entropy’ (*24*, *25*). These metrics were used to identify a rank R that optimally balances interpretability and clustering quality. To ensure

stability, we performed 100 NMF runs at each tested rank using different random seeds and calculated average metric values across these runs.

Sequence motif enrichment was performed on cell type specific ATAC peaks using the findMotifsGenome.pl command from HOMER v4.11 (*26*) with the size flag set to 200 and the JASPAR 2022 database (*27*) obtained from the JASPAR2022 v0.99.7 bioconductor package using the dumpJaspar function from the MonaLisa package. The full set of peaks in a cell type was used as the background set.

### Defining changes in regulatory programs in liver cell types in MASLD

We tested for changes in gene expression, chromatin accessibility, and H3K27ac and H3K27me levels using DESeq2 v1.42.0 (*28*). The cell level count matrix for each modality was split by cell type and counts were aggregated by donor. We performed differential analyses separately comparing MASL, MASH, and MetALD to normal livers using scaled age, scaled BMI, gender, and donor pool as covariates. We also performed differential analyses for fibrosis score as both a quantitative variable as well as a categorical variable to separate MASH/MetALD donors (low:0- 2, high:3-4) including the same covariates, as well as steatosis grade in non-liver disease including fibrosis score as an additional covariate. For each analysis, we excluded donors with<10 cells for the given cell type, features with fewer than 10 total counts across all donors, and categorical contrasts with fewer than three samples in each group. The fGSEA package v1.28.0 was then used to measure enrichment of biological pathways from KEGG (*29*), REACOME (*30*) and GO (*31*) in the differential gene expression results ranked by the ‘stat’ column.

We identified cCREs changing in chromatin state in disease states by classifying chromatin state for each ATAC peak for each disease and fibrosis stage, then performing kmeans clustering of cCREs and manually annotating sites changing in MASH, MASL, or MetALD.

Sequence motif enrichment was calculated in the differentially accessible chromatin sites from the differential cCRE results or within clusters of cCREs using Homer as described above. All tested sites for the given contrast were used as the background set.

Pathway enrichment was calculated for genes linked to cell type specific cCREs using fGSEA with the GO biological processes pathways.

To identify the relationship between gene expression changes in MASL, Fib- MASH, Fib+ MASH, and all MASH livers, we calculated the correlation in effect sizes for each condition compared to normal livers using Spearman’s rho. For cross-modality correlations, we intersected features in each modality in hepatocytes, and calculated Pearson correlation in effect size in Fib+ MASH compared to normal for overlapping features across modalities.

### Gene regulatory network modeling

We identified gene regulatory networks (GRNs) in hepatocytes using SCENIC+(*32*). Seurat SketchData() was employed to down sample either the entire multiome dataset or just hepatocytes in single cell multiome data to 50k nuclei. Utilizing multiome ATAC data from down sampled hepatocytes, we imputed chromatin accessibility and generated binarized topics using pycisTopic. Topics above OTSU calculated thresholds were then retained. Gene expression counts, imputed chromatin accessibility, topics, and differentially accessible peaks in MASLD were then used as input for SCENIC+ (v1.0a1) (*32*). Subsequent downstream analysis was performed with default parameters.

We identified GRNs with increased or decreased activity in MASLD states by performing Gene set enrichment analysis of GRN target genes using the GSEA function of clusterProfiler package, employing the fGSEA method (*33*). GRN information, obtained from SCENIC+ output, was utilized as the TERM2GENE parameter. Selected GRNs and their target genes, based on core enrichment results, were then visualized using Cytoscape(*34*).

### Spatial transcriptomics analysis

Visium HD sequence data was processed using SpaceRanger (v3.0.0) to generate raw count expression matrices with the command “spaceranger count”, using “--loupe-alignment” to manually align the image and the gene expression grid. Bin2cell (v0.3.0) was used to segment the 2x2 micron Visium HD bins into cells, through the StarDist model. StarDist parameters on the H&E image were stardist_model=”2D_versatile_he”, and prob_thresh=0.01, while StarDist parameters used for the gene expression were “stardist_model=”2D_versatile_fluo”, “prob_thresh=0.05”, and “nms_thresh=0.05”. An mpp of 0.25 was used. The counts were then normalized with Scanpy (1.10) using the functions “sc.pp.normalize_total” and “sc.pp.log1p”

Tangram (v1.0.04) was used to map cell type annotations and gene expression information to Visium HD data using cell type markers and gene expression from the RNA portion of the

multiome data. Individual sets of reference cell type markers were generated for different disease groups, namely normal, MASL, Fib-MASH, and Fib+ MASH. To generate cell type markers in each of these groups, cells from the multiome data were subset by each group and then down sampled to 20,000 cells. The Scanpy function sc.tl.rank_genes_group with the parameter “groupby=celltype” was used to identify the top 50 gene markers for each cell type. Tangram models for each spatial dataset was then trained on the cell type markers and gene expression data matching each tissue donor’s respective disease group.

Mapping cell types and gene expression information to each VisiumHD dataset using Tangram first involved splitting the data into patches of 10,000 cells. For each patch, the Tangram function tg.map_cells_to_space with parameters “density_prior=”uniform” and “device=”cuda:0”” was used to find the probability of a particular celltype in a cell. For each patch, the top and bottom 2% quantile probability scores were clipped to remove outliers for each cell type label. The remaining probability scores for each cell type were then min-max scaled, and initial cell type annotations were assigned based on the highest scaled probability score. The separately annotated patches were then merged. Finally, to remove low quality cells, those with a scaled Tangram probability score < 0.5 were removed. To confirm annotations and quantify marker gene expression in spatial profiles, imputed gene expression from Tangram for each Visium HD dataset was down sampled to 100,000 cells. Datasets were then concatenated together, where the average expression of each known marker gene in each cell type was examined.

Hepatocyte zones and sub-cell types were annotated in Visium HD data using Tangram in similar fashion as above. Specifically, Tangram was used to align cell labels and gene expression of hepatocyte subclusters to only those cells within each Visium HD dataset that were previously labeled hepatocytes in the broad cell type Tangram runs. Reference markers for hepatocyte subclusters were identified using the same approach as for broad cell types, using the parameter “groupby=subcelltype” for sc.tl.rank_genes_group to identify hepatocyte subcluster markers. As with the broad annotations, reference markers sets were generated for different disease groups (normal, MASL, Fib-MASH, and Fib+ MASH) with the exception that if a hepatocyte sub-cluster comprised less than 5% of hepatocytes in the multiome dataset from a given disease group, its markers were excluded from the reference marker set for that group.

To plot the expression of specific gene sets or pathways in spatial data, gene module scores for each set were plotted.

### Genome-wide association study enrichments

We utilized genome-wide association (GWAS) data for chronic elevation of alanine aminotransferase (ALT) levels(*35*), a proxy for liver damage and MASLD. We performed GWAS enrichment analyses for cell type-specific regulatory elements using partitioned heritability following LD Score Regression (*36*, *37*). The H3K27ac and ATAC-seq peaks for each liver cell type were lifted to hg19, and annotations were created using the make_annot.py command in LDSC and 1000G EUR reference variants. LD scores were calculated using the ldsc.py command with 1000G_EUR_Phase3_baseline_snps and a ld window of 1cM. Finally, enrichment was calculated using the ldsc.py command with the --h2-cts flag, with the 1000G_EUR_Phase3_baseline and all peaks of a given modality as background sets in the analysis. The same analysis was run on ATAC-seq peak sites split by chromatin state using the total set of sites for that cell type in the background set. All enrichments were used for calculating FDR using Storey’s method in the package FDR v2.34.0, and tests were considered significant at FDR<.10.

We performed enrichment analyses of subsets of sites with altered activity in MASLD for fine-mapped variants using FINRICH v0.3.2, which performs a permutation test to identify enrichment of credible set variants in genomic windows relative to a background set. For differential ATAC sites all features passing 5% FDR were used as the foreground and all tested sites the background against credible set variants from the cALT GWAS trans-ancestry analysis lifted over to hg38 using ‘SNPlocs.Hsapiens.dbSNP155.GRCh38’ v0.99.24. Fold-enrichment was assessed for sites changing in chromatin state and for regions from GRNs changing in activity in MASL and MASH from SCENIC+. The significance of the fold-enrichments was determined via permutations followed by FDR correction. We identified credible set variants from the cALT trans-ancestry fine-mapping overlapping cCREs in GRNs for each TF and constructed a graph of TF GRNs based on credible set variant overlap using igraph v2.1.4.

### Quantitative trait locus mapping

We performed QTL mapping using the RNA, ATAC, H3K27ac, and H3K27me3 modalities. Barcode level count matrices were subset by cell type and counts were aggregated by donor. The resulting libraries were transcripts per million (TPM) normalized (RNA) or counts per million (CPM) normalized and features were filtered based on at least 20% of samples with >0.1 TPM or CPM and 6 counts. The counts were normalized using a TMM adjustment in edgeR v4.0.16 (*38*) and

inverse-normal transformed. Count matrices were converted to bed format as described on the TensorQTL GitHub repository (https://github.com/broadinstitute/tensorqtl). For ATAC and DPT modalities the bed coordinates were the feature peak coordinates, and for RNA TSS sites were derived from the GTF used in CellRanger based on strand and start and end coordinates, with the bed coordinates being a 1bp region at the start of the gene.

We generated 10 genotype PCs based on analysis in plink v1.9 to account for ancestry. Sample genotypes were merged with 1000 Genomes genotypes and pruned prior to PCA. To account for latent effects in genomic profiles we ran PCA on the inverse-normal transformed count matrices and extracted the first 15 PCs. We used gender, scaled age, scaled BMI, fibrosis stage, disease label, and sequencing pool as additional biological and technical covariates.

TensorQTL v1 (*39*) was run on a HPC at the San Diego Supercomputer Center with an a100 gpu. First, pairwise summary stats were generated using the --cis_nominal mode, then gene wise permutations and FDR cutoffs were determined using the --cis mode with the --fdr 0.05 flag, which performs permutations for each gene using the lead SNP to determine an empirical p-value and performs Storey’s method for FDR correction on the permuted p-values. These empirical p-values are back mapped to determine the nominal p-value cutoff for other variants for the given feature, which were back mapped onto the pairwise summary stats.

We followed the mashr v0.2.79 (*40*) eQTL analysis vignette for each modality independently. First, we constructed two mashr datasets, one comprising the beta and error terms of the summary stats for 200,000 randomly selected feature variant pairs, and the other representing every feature-variant pair significant in any cell type. The randomly selected dataset was used to estimate the canonical covariance matrix, and the true effects the data drive covariance matrix. These two matrices and the randomly selected dataset were used to train the mash model. Then the model was used to estimate the effects in the dataset of significant associations using the get_fitted_g parameter. QTLs determined as specific based on a lfsr value below 0.05.

Summary stats for Vujkovic et al 2022 (*35*) were accessed from dbGaP and converted to hg38 coordinates using SNPlocs.Hsapiens.dbSNP155.GRCh38 in Bioconductor. Any QTL feature passing 5% FDR was colocalized with GWAS summary statistics using the coloc.abf function from coloc v5.2.3 (*41*). Signals were determined to be colocalized based on PPH4 greater than 0.8, the GWAS signal passing genome wide significance (5x10^-8^), and the lead GWAS and QTL variants being in high LD (r^2^>0.5).

Cross-modality QTL modules were created following the process outlined in Arthur et al (*42*). We identified QTL features by cell type with overlapping *cis* QTL mapping windows and performing colocalization. Colocalization signals passing PPH4>0.8 were constructed into a connected graph using igraph v2 (*43*) and clustered using the Louvain algorithm. Clusters with modularity scores above 0.3 were divided into a subgraph and clustered recursively.

### Sequence-based models of chromatin accessibility

A ChromBPNet model was trained using the default preprocessing parameters outlined in the ChromBPNet tutorial (https://github.com/kundajelab/chrombpnet/wiki/Preprocessing). To account for Tn5 bias, a separate model was trained using our own 10x multiome pseudobulk data. Four bias-factorized models were trained on hepatocytes under different conditions: Control, MASL, MASH, and MetALD. One model was trained using untreated hepatocytes, while the other three were trained using hepatocytes from MASH, MASL, and MetALD conditions. For training, we used macs2 narrowPeak regions with a p-value threshold of <0.05, called on pseudobulk ATAC fragments from all hepatocyte samples. Chromosomes 1, 3, and 6 were assigned for training, while chromosomes 8 and 20 were reserved for validation. Following training, default parameters were applied. ChromBPNet models consist of two output heads: (1) the profile head, which predicts the shape of a profile, and (2) the counts head, which estimates the total counts within a profile. The chrombpnet_nobias.h5 models were used for this analysis, and we computed sequence contribution scores for one or both heads. Additionally, we utilized ChromBPNet for variant prediction to assess the impact of genetic variants on chromatin accessibility within our dataset. We considered variants where the absolute value of log_counts_diff was greater than .025 as having a functional effect. To generate predicted counts and contribution bigwigs for both alleles of example variants we used bcftools to insert the alternate allele into the reference bigwig and ran the pred_bw and contri_bw functions from ChromBPNet on both the standard and custom reference genome using the chrombpnet_nobias.h5 model.

### Annotating MASLD associated loci in hepatocytes

We identified loci associated with MASLD (cALT levels) from a large genetic association study (*35*). We identified loci where the lead fine-mapped variant was a significant QTL in hepatocytes for chromatin accessibility, H3K27ac or H3K27me3 histone modifications, or gene expression levels, as well as loci that were statistically colocalized with hepatocyte QTL association (shared probability>.8). We next identified loci where at least one fine-mapped variant overlapped a hepatocyte cCRE or active chromatin state, as well as loci where at least one fine-mapped variant had a predicted effect on hepatocyte chromatin accessibility from chromBPNet models. Finally, we annotated target genes that were either within 10kb of a chromatin loop to a hepatocyte cCRE harboring fine-mapped variants or a colocalized eQTL.

### Identification of trajectories and pseudotime

To infer potential state changes between hepatocyte subtypes during we performed trajectory analysis using the Monocle3 package (v1.3.4). Briefly, we selected all hepatocyte cells from high fibrosis donors (MASH/MetALD with fibrosis score >=3) and learned a trajectory with monocle’s learn_graph() function with the following parameter settings: maximum euclidean distance ratio of 1, minimal branch length of 10, 25 nearest neighbors, and the default method for identifying nearest neighbors. Next, to reconstruct pseudotime from the Hepatocytes.Injured cluster, we chose the node furthest in that cluster as a starting point and ordered all cells from there. The top 50 genes with the largest fold change in Fib-a hepatocytes when comparing high fibrosis (Fib+) MASH against normal livers were plotted along the trajectory after aggregating cells into bins (n=50) based on pseudotime order.

### Identification of Fibrosis-adjacent hepatocytes and neighborhood enrichment analysis

We first generated a fibrosis-associated gene set by selecting the top 50 genes with the largest fold change in the fibrosis-associated hepatocytes when comparing high fibrosis (Fib+) MASH against normal livers. These genes also mark a subset of cells identified in multiome pseudotime analysis described below. Module scores for this fibrosis-associated gene set were then calculated for each hepatocyte in a Fib+ MASH Visium HD dataset (HL2201019). Hepatocytes in the top 1% of module scores (>0.2) were isolated and labeled "Fib-adj hepatocytes" and used this population was used for neighborhood enrichment analysis.

Neighborhood enrichment was calculated using the sq.gr.nhood_enrichment function from Squidpy (c1.6.1) (sq.gr.nhood_enrichment (adge_comb, cluster_key="subcluster_label_comb", show_progress_bar=False)). Enrichment was evaluated based on combined broad cell type classifications for non-hepatocytes, hepatocyte sub-cluster annotations, and Fib-adj hepatocytes.

### Cell-cell interaction analyses

We performed cell-cell communication analysis on Visium HD spatial transcriptomic data using CellChat v2.1.0. A CellChat object was created from count data and meta data including Tangram cell types using the createCellChat function where spatial.factors were calculated with a conversion factor of 0.5 and a spot size of 2. Over expressed genes and interactions were calculated using the identifyOverExpressedGenes and identifyOverExpressedInteractions functions. Interaction probabilities of all L-R pairs in the cellchat database were calculated using the computeCommunProb function, employing the truncatedMean method with the following parameters: trim = 0.05, distance.use = TRUE, interaction.range = 50, scale.distance = 1, contact.dependent = TRUE, and contact.range = 15. The resulting CellChat object was filtered to retain interactions involving a minimum of 10 cells expressing either the ligand or receptor. These interactions were then converted into a data frame using the subsetCommunication function. To account for multiple comparisons, we applied the Benjamini-Hochberg method via the q-value package, considering predicted interactions with an FDR < 0.1 as significant. We generated a heatmap of weighted interactions using the netVisual_heatmap function in CellChat and additional visualizations were constructed using the ggplot2 package.

## Supplementary Files

### Supplementary Tables

Table 1 – Human liver donors profiled in study

Table 2 – Marker genes to annotate liver cell types

Table 3 – Changes in liver cell type and sub-type abundance in MASLD

Table 4 – Biological pathways enriched in TF GRNs

Table 5 – Fine-mapped MASLD variants annotated in liver cell types

Table 6 – Cell type-specific cCREs and target genes in the liver

Table 7 – Biological pathways enriched in genes associated with MASLD

Table 8 – Predicted target genes of hepatocyte cCREs altered in MASLD

Table 9 - Sequence motifs enriched in cCREs altered in MASLD

Table 10 – Sequence motifs enriched in hepatocyte cCRE modules altered in MASLD

Table 11 – TF GRNs with altered activity in MASLD

Table 12 – Liver cell type QTLs

Table 13 – Sequence motifs enriched in liver cell type QTLs

Table 14 – Liver cell type QTLs at MASLD loci

Table 15 – Marker genes and cCREs of hepatocyte sub-clusters

Table 16 – Motifs enriched in hepatocyte sub-cluster specific cCREs

Table 17 - Pathways enriched in hepatocyte sub-cluster genes altered in MASLD

Table 18 – Motifs in hepatocyte sub-cluster cCREs altered in MASLD

### Supplementary Data

Liver cell type-specific cCREs

Liver cell type-specific chromatin states

Liver cell type-specific TF gene regulatory networks Liver cell type-specific chromatin loops

Differential association results for molecular features in liver cell types Summary statistics for liver cell type QTLs

The supplementary Data, due to the large file sizes (>500MB), has been deposited to a public database Zenodo (record ID: 15298484).

---

**Supplementary Figure 1.**
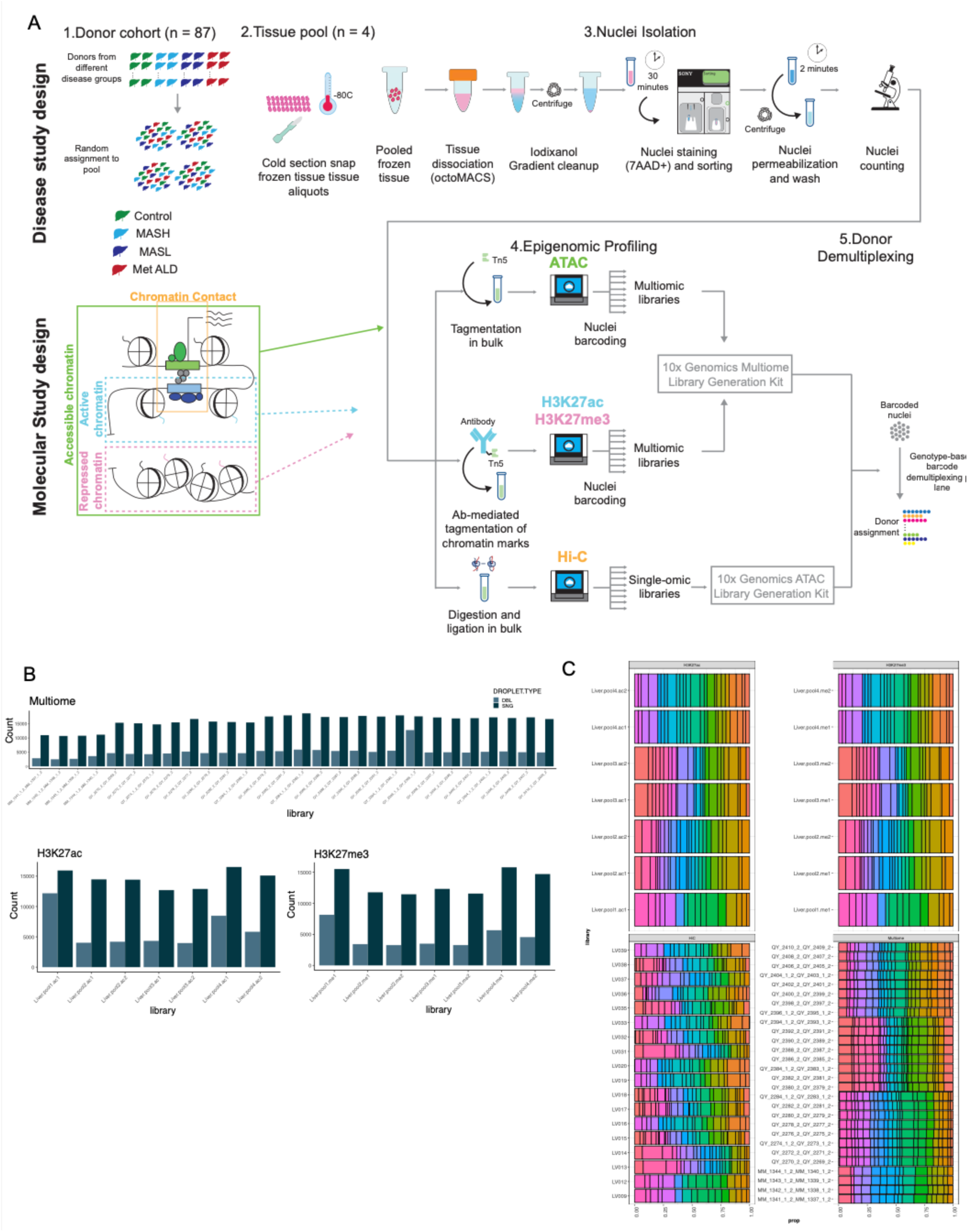
Study design for single cell profiling of human livers. A) Design of study to perform single cell assays of human livers in sample pools. Livers from normal and MASLD groups (MASL, MASH, MetALD) were randomly assigned to groups. Samples from livers in each group were pooled, and then nuclei were extracted from each pool. Multiple single cell libraries including 10x multiome, H3K27ac and H3K27me3 Paired-Tag and Droplet Hi-C were generated on nuclei from each pool, and libraries were sequenced. Finally, the donor of origin was assigned to barcodes in each pool by demultiplexing by using genotyping array data for each donor. B) Number of singlet and doublet barcodes obtained from demultiplexing of libraries for each single cell assay. C) Proportion of cells from each liver donor obtained from libraries for each single cell assay.

**Supplementary Figure 2.**
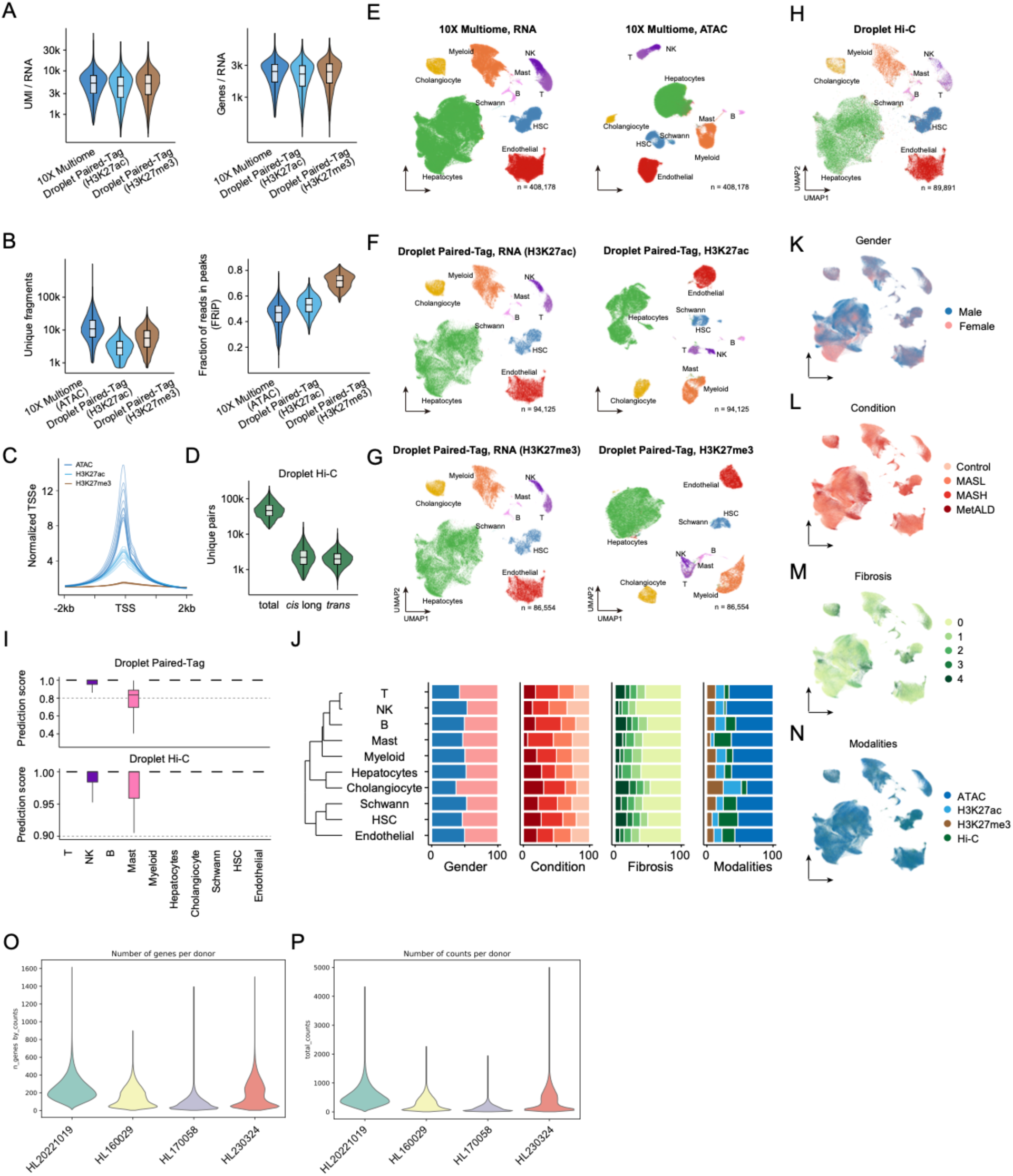
Characteristics of single cell profiles in liver. A) Number of UMIs (left) and genes (right) per barcode for the RNA modalities for 10x Multiome and Droplet Paired-Tag as counted by Seurat. B) Fragment length (right) and fraction of reads in peaks (left) for ATAC and DNA modalities of Droplet Paired-Tag. C) Enrichment of normalized ATAC, H3K27ac, and H3K27me3 signal around TSS sites (+-2kb) per single cell library. D) Unique read pairs in Droplet Hi-C including *cis* long and *trans* contacts. E) 10X Multiome UMAP and cell type annotations. F) H3K27ac Droplet Paired-Tag UMAP and cell type annotations. G) H3K27me3 Droplet Paired-Tag UMAP and cell type annotations. H) Droplet Hi-C UMAP and cell type annotations. I) Label transfer scored for Droplet Paired-Tag and Hi-C by cell type. J) Proportion of disease, gender, and modality composition across cell types, visualized in UMAP space in K-N. O) Number of detected genes per donor after filtering in spatial transcriptomics profiling of human livers. P) Number of counts per donor after filtering in spatial transcriptomics profiling of human livers.

**Supplementary Figure 3.**
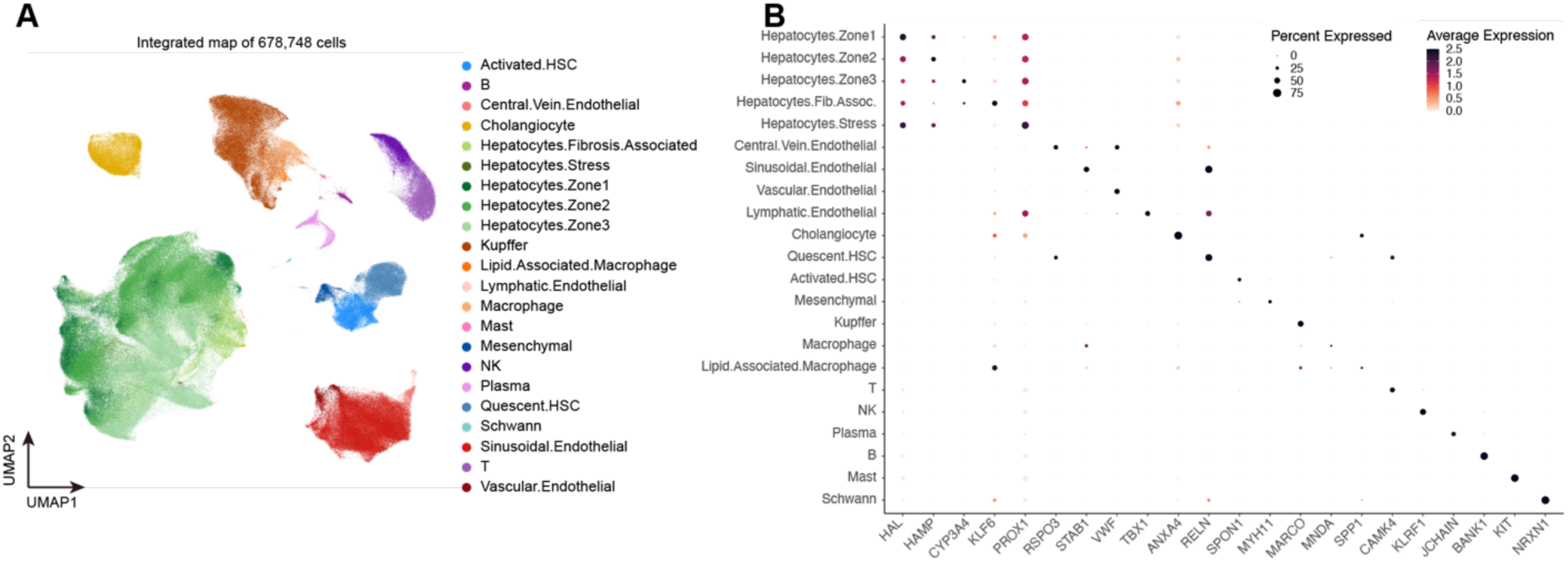
Identifying sub-types of liver cell types in single cell profiles. A) UMAP of combined 10X Multiome, Droplet Paired-Tag and Droplet HiC data showing cell sub- type definitions in the liver. B) Expression of selected marker genes for hepatocyte sub-types.

**Supplementary Figure 4.**
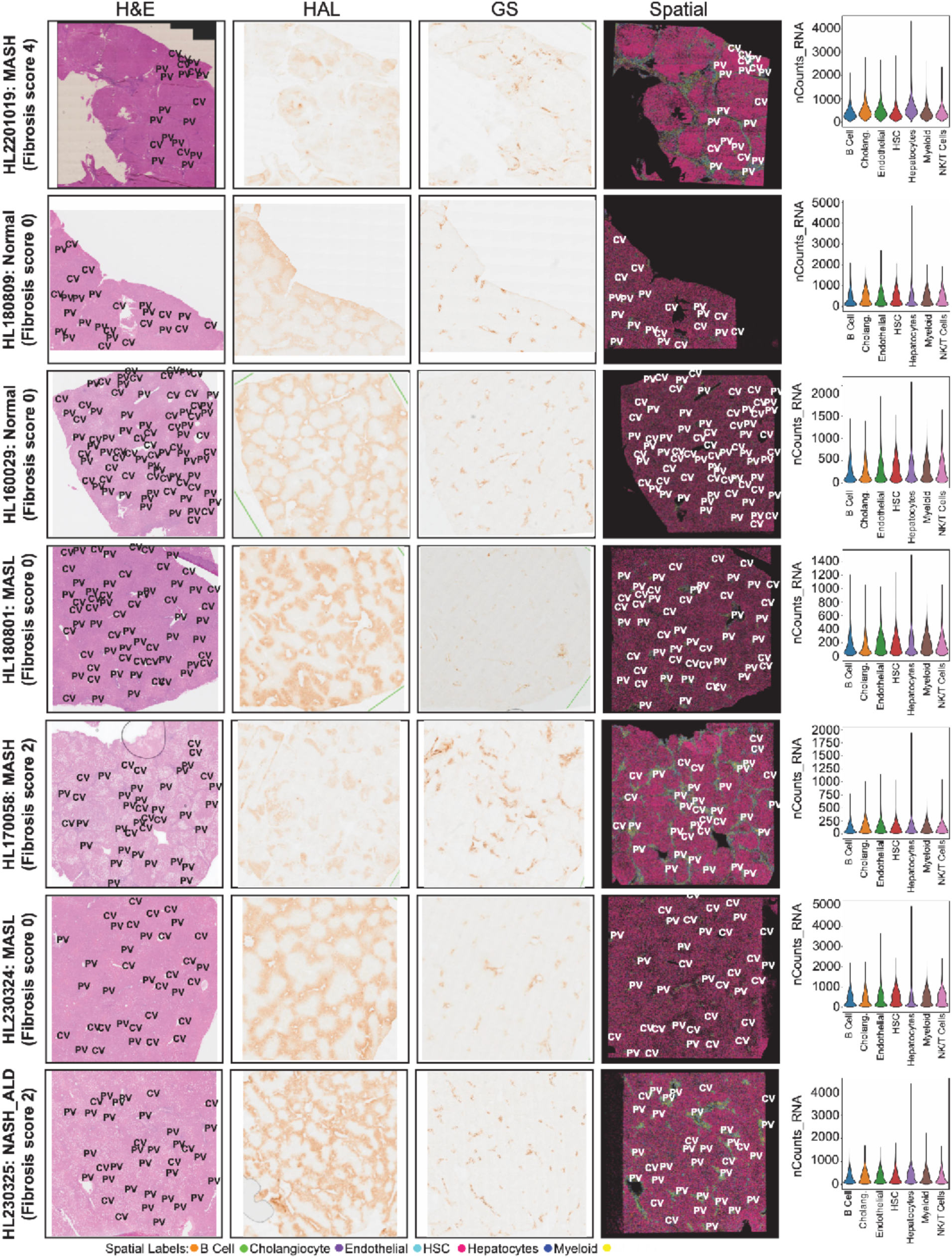
Histological, immunohistochemical, and spatial transcriptomic profiling across MASLD disease stages. Liver sections from seven donors across MASLD, including normal, MASL, MASH (Fibrosis score 0–4), and MetALD—were profiled using histology (H&E), immunohistochemistry (HAL and GAS staining), and 10x Visium HD spatial transcriptomics. Each row corresponds to an individual liver sample (donor ID indicated at left with fibrosis stage noted). Left panels show H&E staining with annotated central vein (CV) and portal vein (PV) regions. Middle panels display HAL and GAS staining marking periportal and pericentral regions, respectively, and highlighting histological variation across conditions. Spatial transcriptomic maps (fourth column) show the spatial distribution of six major cell types (hepatocytes, cholangiocytes, endothelial cells, HSCs, myeloid cells, and B cells), inferred via Tangram mapping. Rightmost panels show violin plots of RNA counts per cell type across each sample. Notably, the severe fibrosis MASH sample (top row) shows architectural disruption and altered spatial composition compared to normal and MASL livers.

**Supplementary Figure 5.**
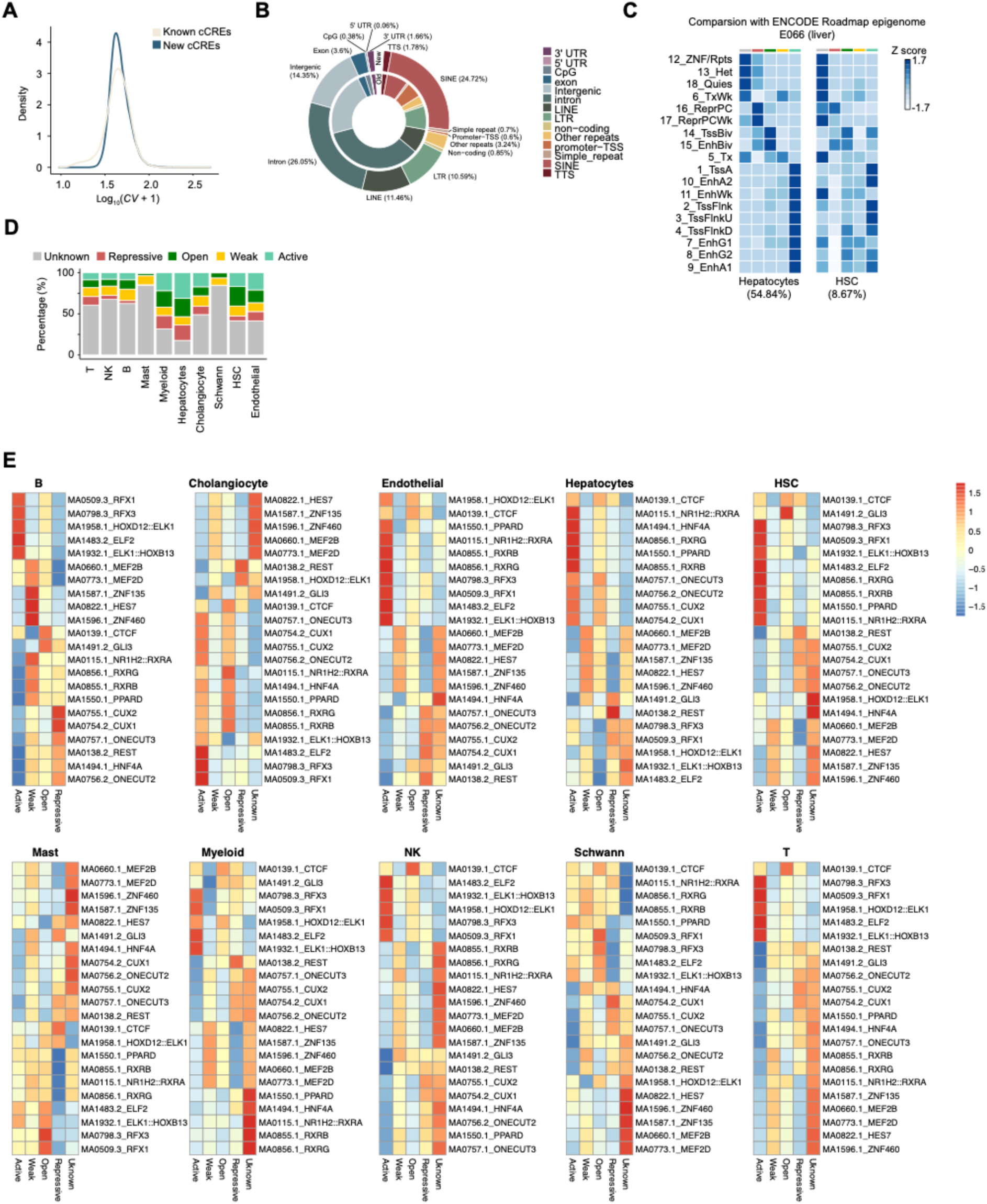
**Characteristics of liver cell type cCREs**. A) Variability (coefficient of variation) in chromatin accessibility signal (CPM) of novel cCREs identified in this study compared to known cCREs. B) Distribution of known and novel cREs across genomic features. C) Overlap of chromatin state annotations in hepatocytes and HSCs to chromatin states in bulk liver from the Epigenome Roadmap. Parenthesis show the percentage of each cell type among all cells in the single cell maps. D) Proportion of all cCREs classified in each chromatin state for each liver cell type. E) TF sequence motifs enriched in each chromatin state for each liver cell type using Homer.

**Supplementary Figure 6.**
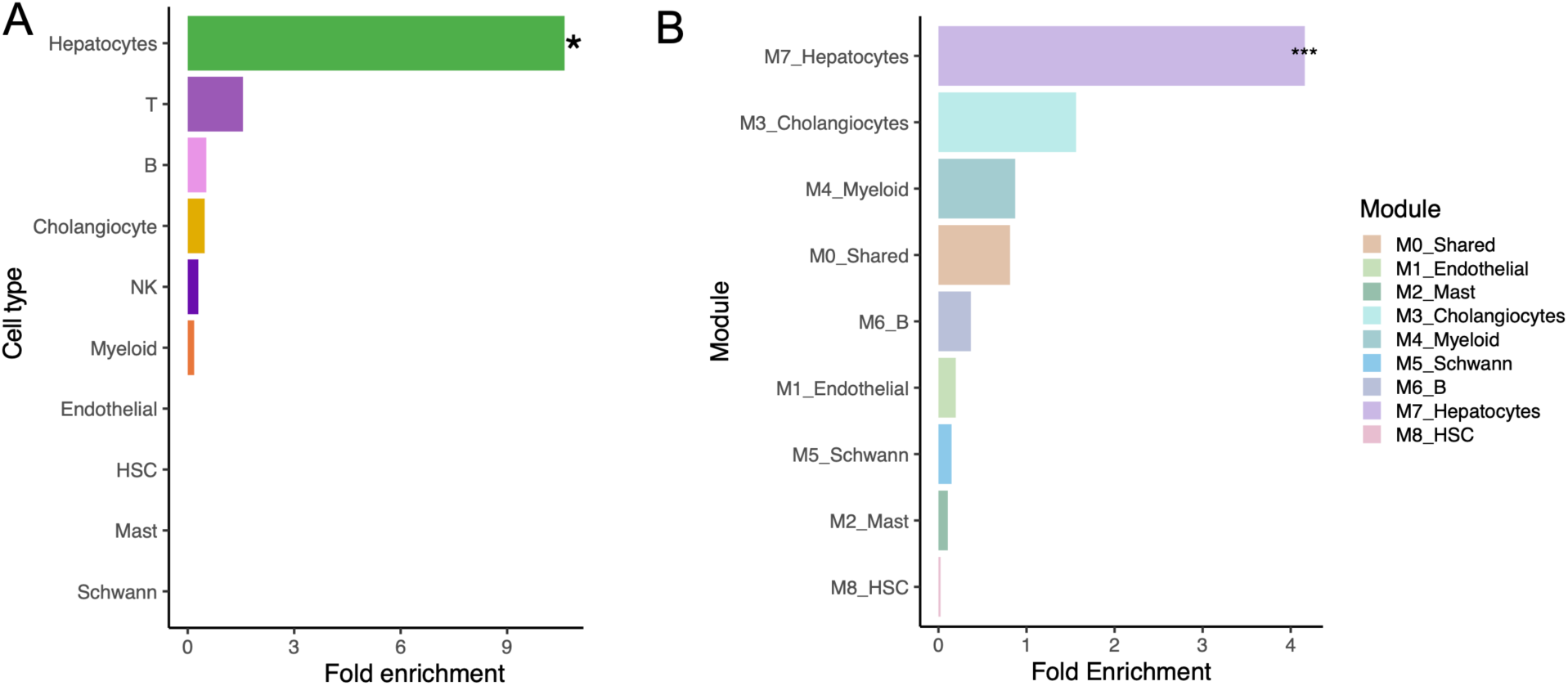
Enrichment of MASLD variants for cell type-specific cCREs. Enrichment of fine-mapped variants for cALT/MASLD proxy GWAS in (A) liver cell type-specific cREs and (B) Liver cell type cCREs within cCRE modules (M0-9)

**Supplementary Figure 7.**
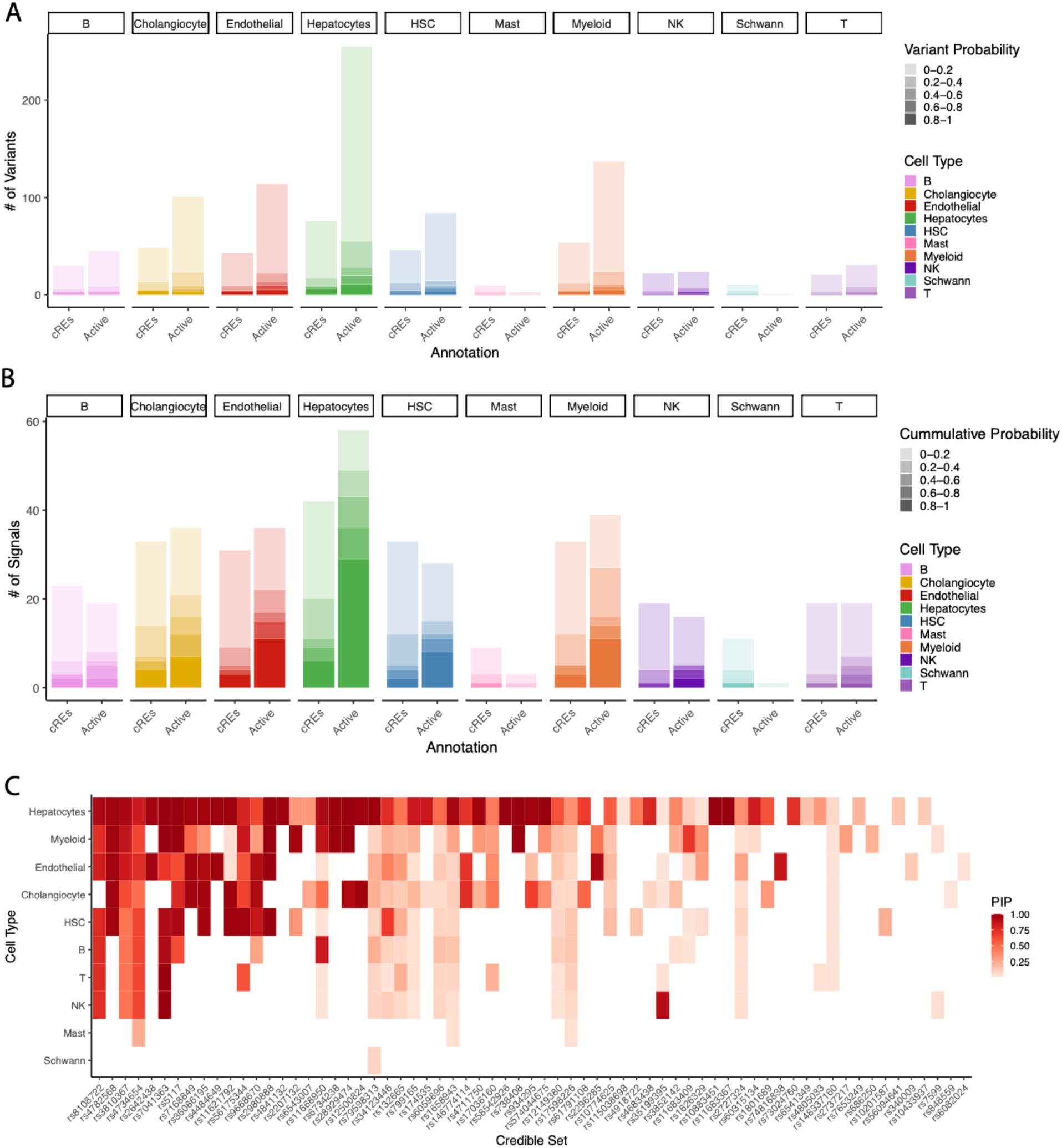
MASLD variants annotated in liver cell type cCREs. A-B) Number of fine-mapped variants (A) and credible sets (B) cALT/MASLD proxy GWAS variants intersecting liver cell type cREs and active chromatin state regions, stratified by variant posterior inclusion probability (PIP) C) Annotation of specific fine-mapped signals to liver cell types by intersection with cREs, colored by probability of variants annotated.

**Supplementary Figure 8.**
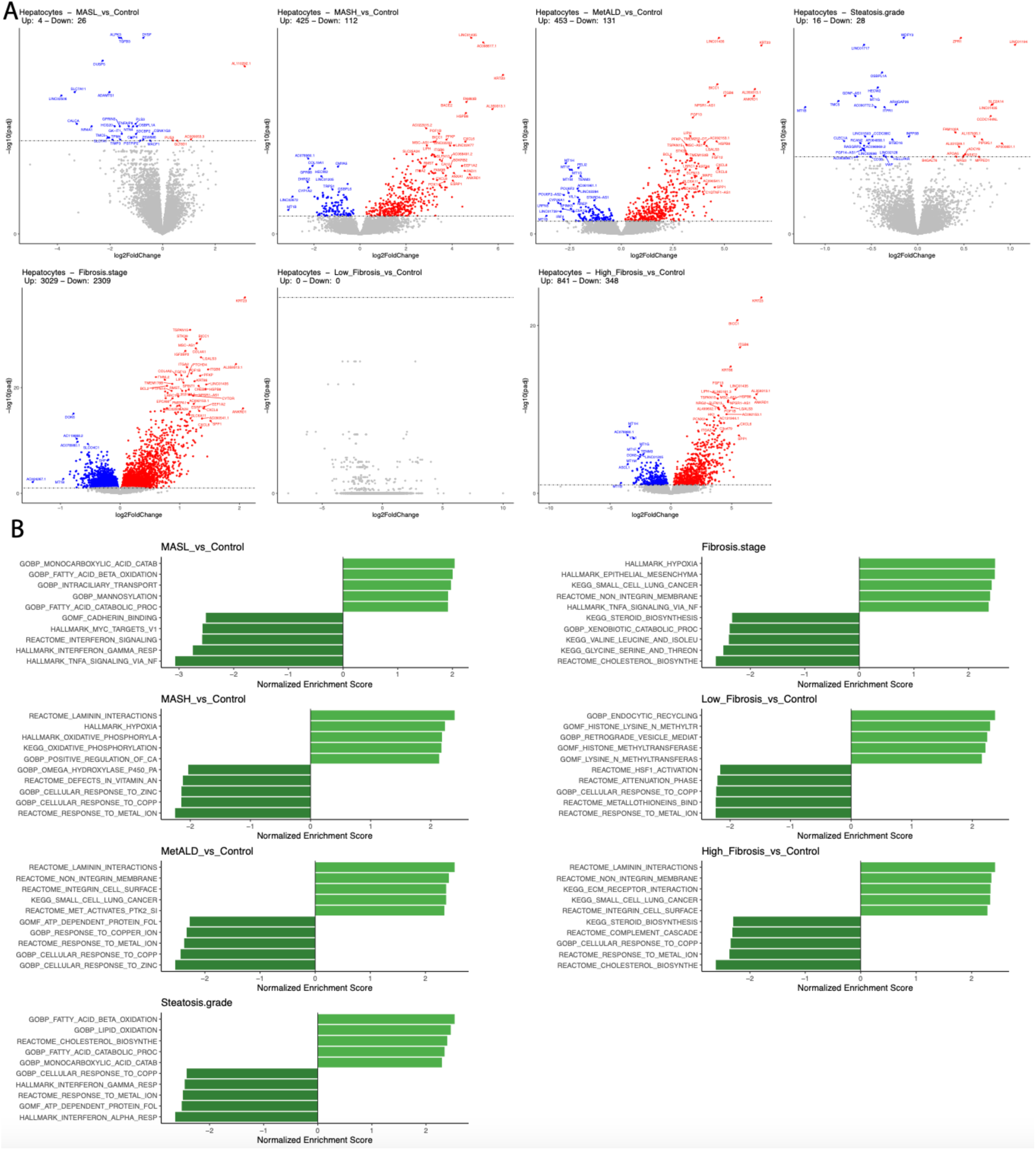
Changes in gene expression in hepatocytes in MASLD. A) Differentially expressed genes from DESeq using the 10X Multiome data for listed contrasts. Red dots highlight upregulated genes in disease, blue dots highlight down regulated genes, and grey represents genes not significant at 10% FDR. B) Pathway enrichments for differentially expressed genes using fGSEA. The top 5 most significantly enriched and depleted pathways are plotted.

**Supplementary Figure 9.**
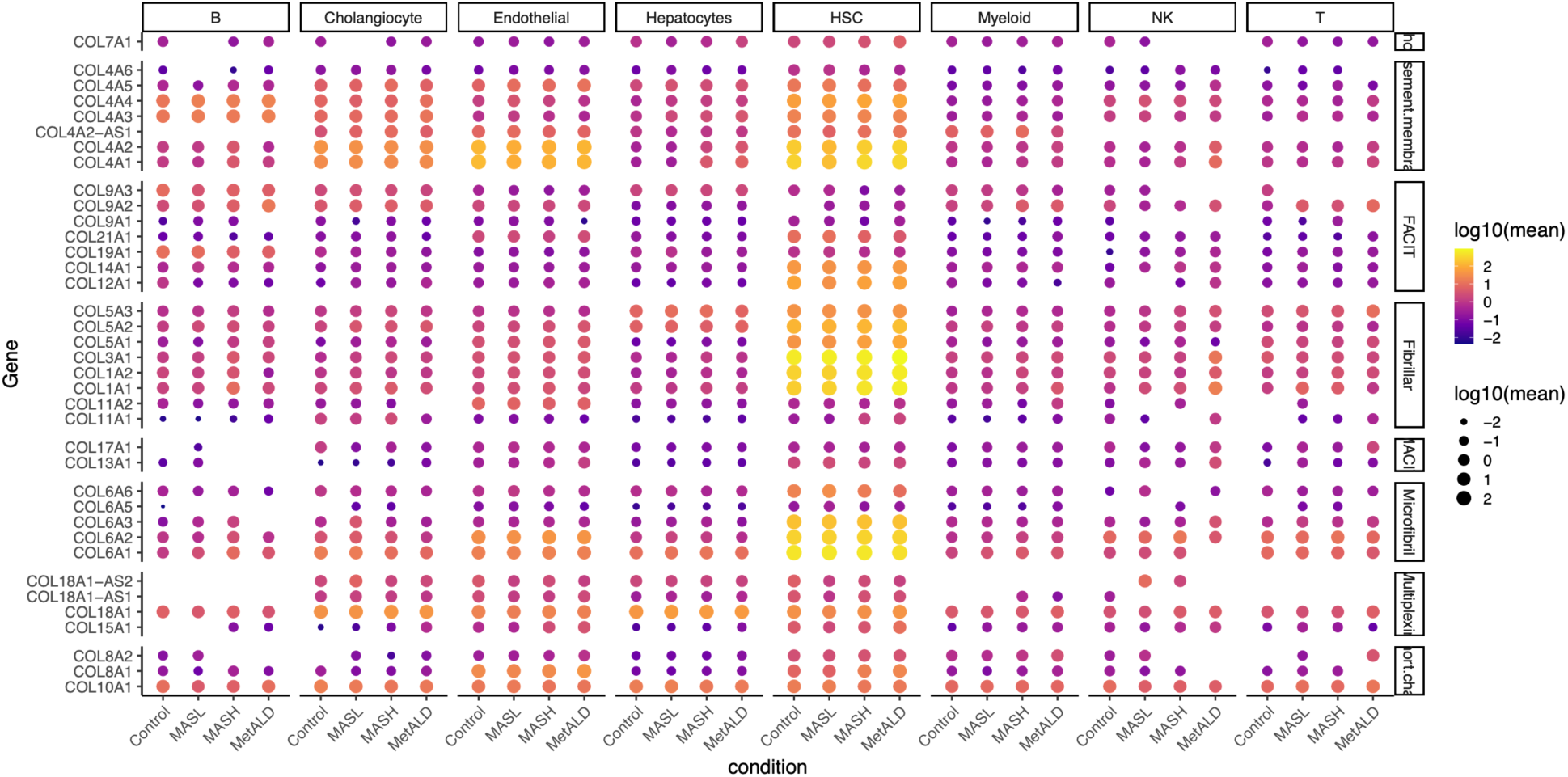
Change in collagen family gene expression in MASLD progression. A) Log-transformed average expression of genes in the collagen family in liver cell types across MASLD stages

**Supplementary Figure 10.**
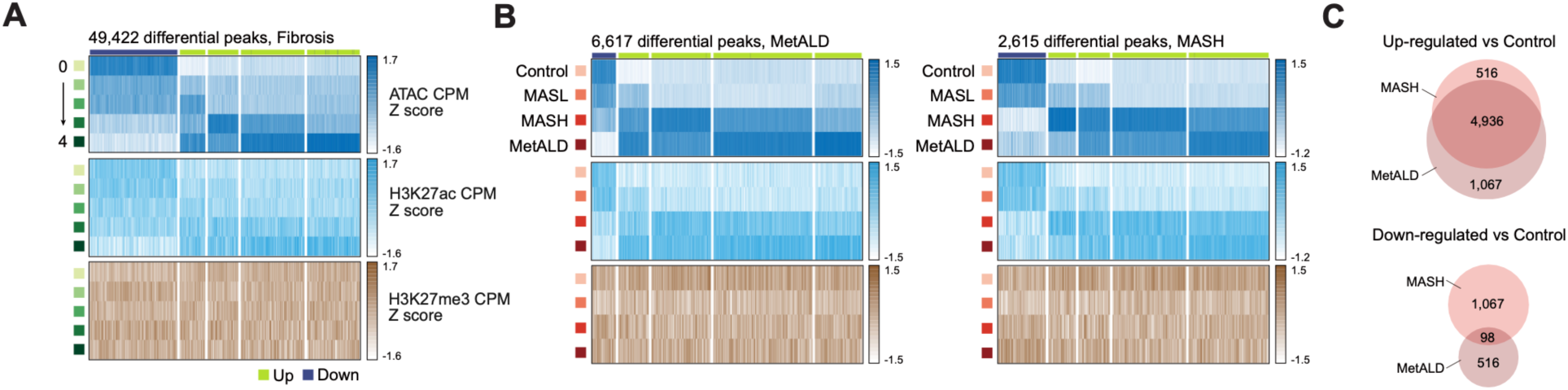
Changes in hepatocyte cCRE activity in MASLD. A) Differentially accessible cCREs (ATAC) across fibrosis stages clustered based on the epigenomic signal at different fibrosis stages. ATAC signal in navy, H3K27ac signal in blue, and H3K27me3 signal in brown is shown for each site. B) Clustering of differentially accessible cCREs in MetALD versus Control and MASH versus Control, as in A. C) Overlap of cCREs changing in MASH and MetALD.

**Supplementary Figure 11.**
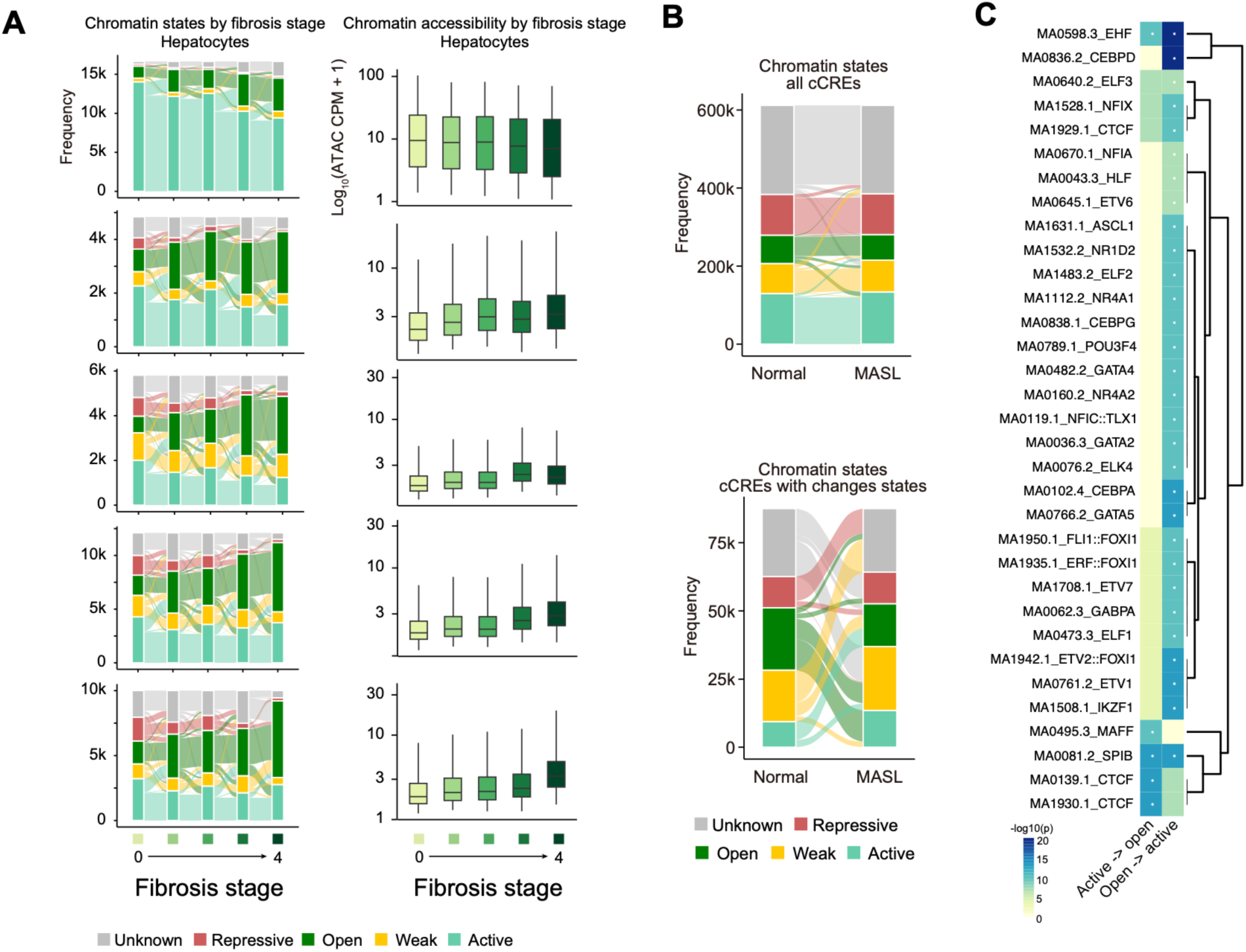
Changes in hepatocyte chromatin state in progression to MASH. A) Chromatin state changes across fibrosis stage within clusters of hepatocyte cCREs associated with fibrosis stage (left) and average chromatin accessibility (CPM) across fibrosis stages for the same cCREs (right). B) Chromatin state in normal and MASL livers across all hepatocyte cCREs (left) and only hepatocyte cCREs that change state between normal and MASL livers (right). C) Sequence motif enrichment in cCREs that transition from open state in normal to active state in MASL, or from active state in normal to open state in MASL.

**Supplementary Figure 12.**
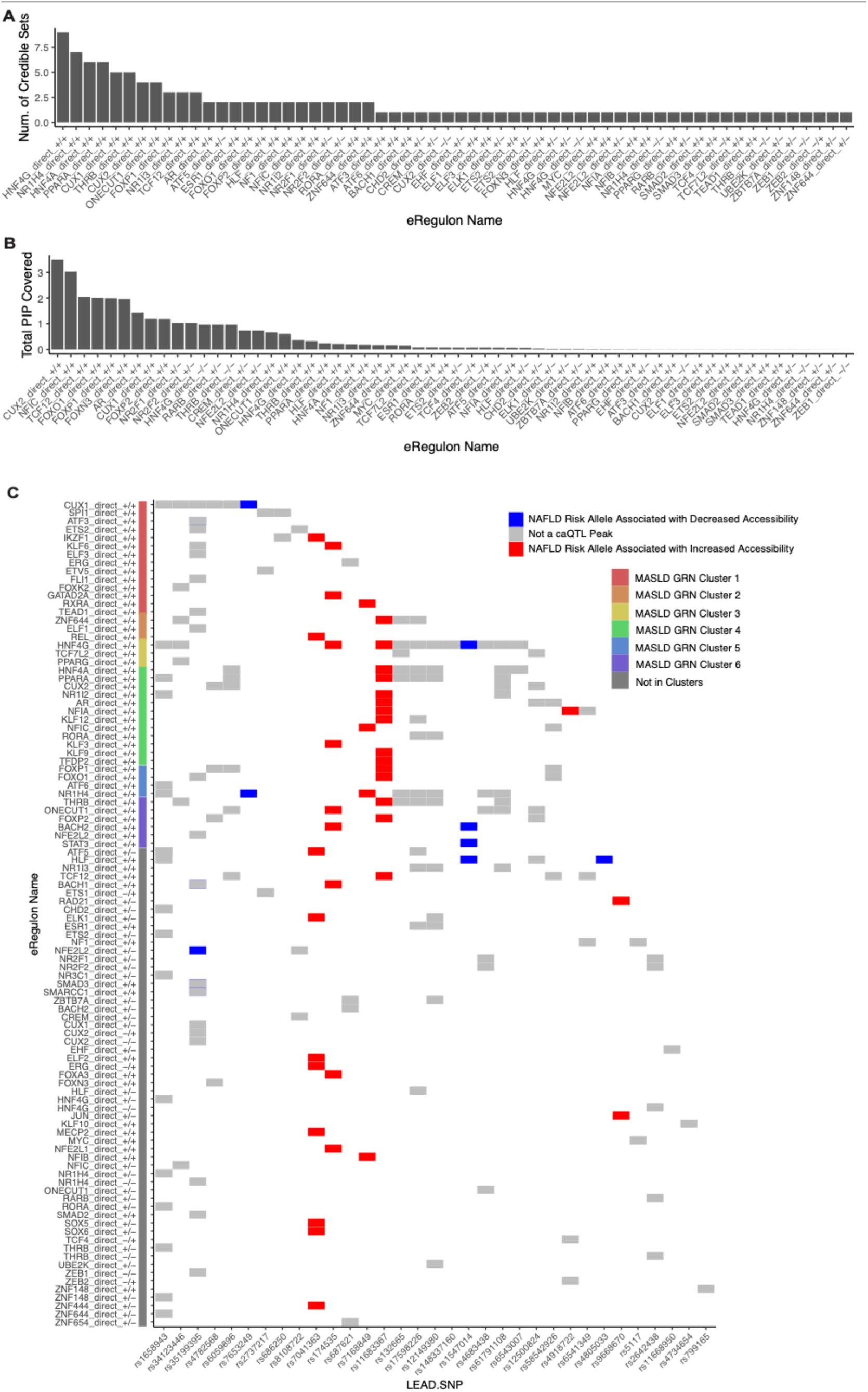
Enrichment of MASLD variants in TF GRNs. A-B) Number of credible sets overlapping GRN cCREs and cumulative PIP of variants overlapping GRN cCREs. C) Break down of variants annotating to GRN cCREs and the direction of accessibility relative to the risk allele in QTL signals. Red is increased accessibility with the risk allele, blue is decreased accessibility with the risk allele, and grey is no QTL association. GRNs are colored by cluster.

**Supplementary Figure 13.**
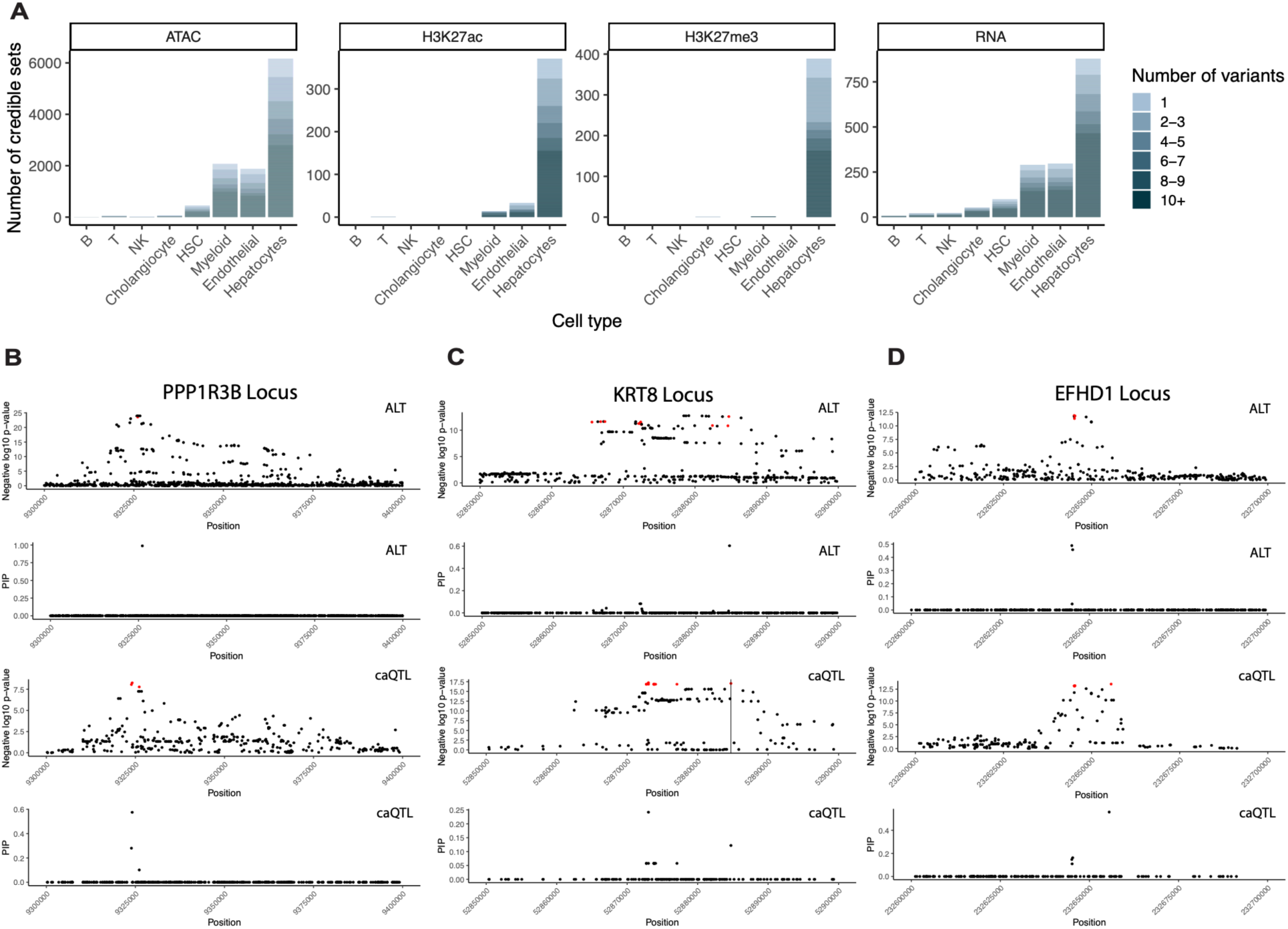
Fine-mapping of liver cell type QTLs. A) Summary of the number of fine-mapped QTL signals (credible sets) for each modality and the number of credible set variants per signal. B-D) Locus zoom plot for the MASLD proxy GWAS signal (top), variant pips for GWAS fine-mapping (upper middle), caQTL locus zoom plot (lower middle), and variant pips from caQTL fine mapping (bottom) for the *PPP1R3B* (B), *KRT8* (C), and *EFHD1* (D) loci.

**Supplementary Figure 14.**
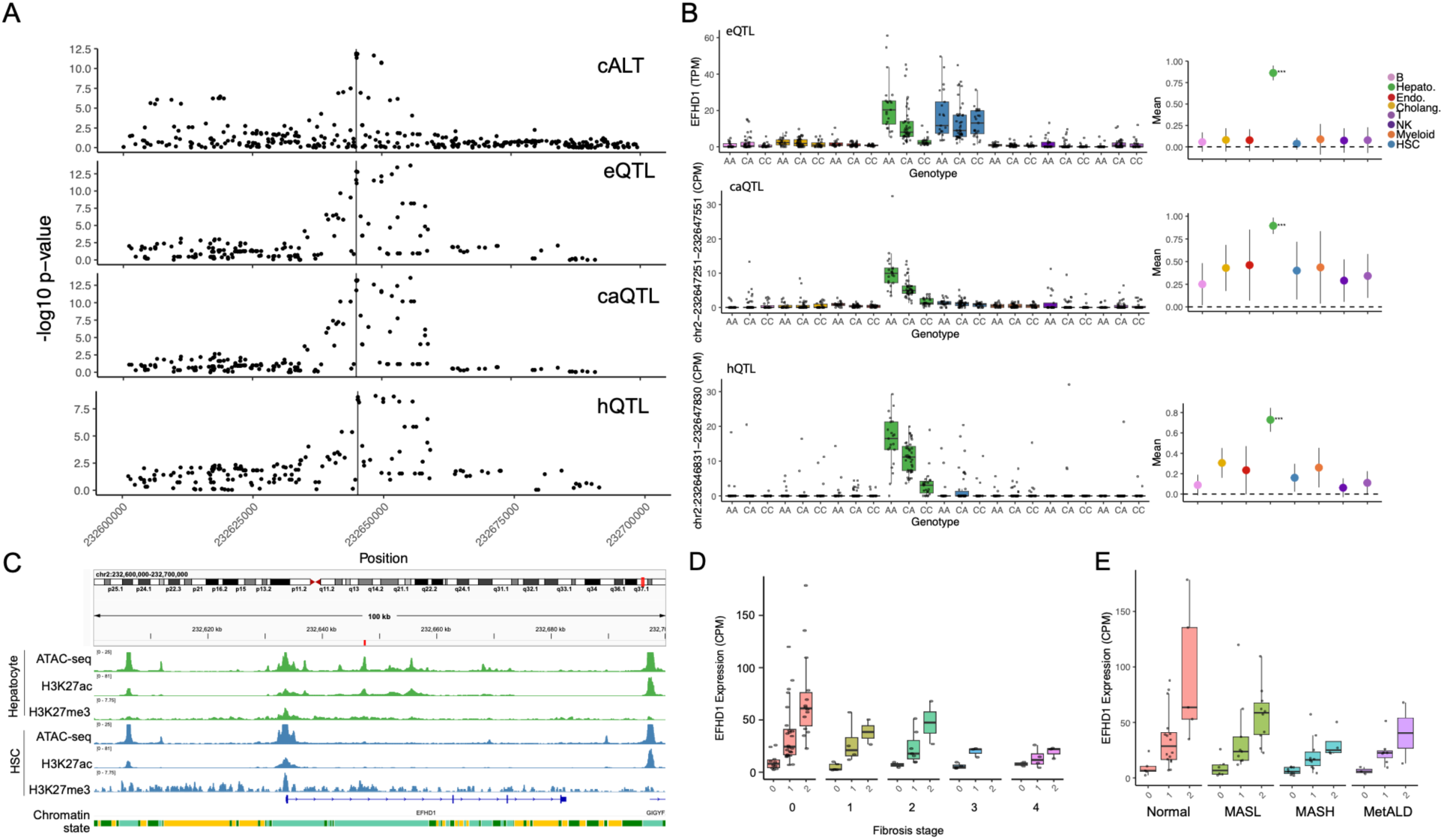
MASLD risk variants affect *EFHD1* expression. A) eQTL (top), caQTL (middle) and H3K27ac hQTL (bottom) boxplots and mashr effect estimates for the *EFHD1* locus. The vertical line represents the lead fine-mapped MASLD proxy SNP. B) Mashr estimates for the lead fine-mapped MASLD variant including the estimated effect and standard error, where asterisks for local false sign rate (LFSR) p-values are: ****0.0001, ***0.001, ** 0.01, and *0.05. C) Locus zoom plots for MASLD proxy GWAS (top), EFHD1 eQTL (upper middle), caQTL (lower middle), and H3K27ac hQTL (bottom). D,E) Interaction plot of effect between *EFHD1* expression and fibrosis stage (E, interaction term p-value = 0.046) and MASLD stage (F, interaction term p-value = 0.049).

**Supplementary Figure 15.**
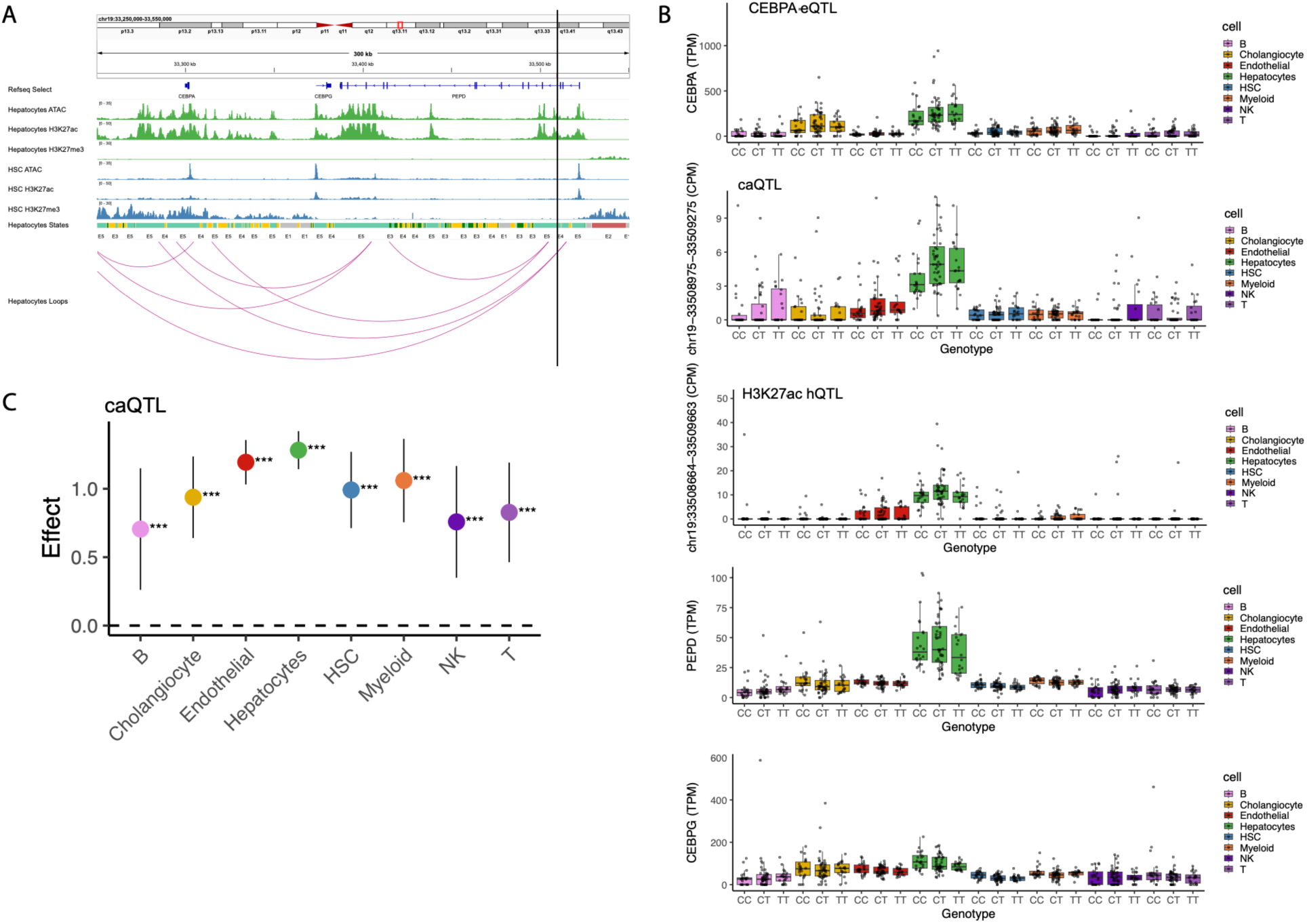
MASLD risk variants interact with distal target gene *CEBPA* at the 19q13 locus. A) Browser track of the 19q13 locus showing ATAC, H3K27ac, and H3K27me3 signal in hepatocytes and HSCs, and chromatin states and chromatin loops in hepatocytes. The vertical black bar marks the MASLD-associated caQTL peak which interacts with *CEBPA*. B) *CEBPA* eQTL boxplot (top) and caQTL (middle) and H3K27ac hQTL (bottom) for the linked cRE. The bottom plots show eQTL boxplots for *PEPD* (top) and *CEBPG* (bottom), the two other genes at the locus. Neither *PEPD* or *CEBPG* had significant eQTL association. C) mashr estimates for the caQTL at this locus. Asterisks for local false sign rate (LFSR) p-values are: ****0.0001, ***0.001, **0.01, and *0.05

